# Polygenic susceptibility to dilated cardiomyopathy underlies peripartum, alcohol-induced, and cancer therapy-related cardiomyopathies

**DOI:** 10.1101/2025.02.17.25321705

**Authors:** Dimitri J. Maamari, Kiran J. Biddinger, Sean J. Jurgens, Joel T. Rämö, Liam Gaziano, Alice Zheng, Dolphurs Hayes, Carlos A. Gongora, FinnGen, Seung Hoan Choi, Zoltan Arany, Paaladinesh Thavendiranathan, Akl C. Fahed, Amy A. Sarma, Tomas G. Neilan, Amit V. Khera, Patrick T. Ellinor, Krishna G. Aragam

**Affiliations:** Cardiovascular Disease Initiative, Broad Institute of MIT and Harvard, Cambridge MA, USA; Department of Medicine, University of Texas Southwestern Medical Center, Dallas, TX; Cardiovascular Research Center, Massachusetts General Hospital, Harvard Medical School, Boston MA, USA; Center for Genomic Medicine, Department of Medicine, Massachusetts General Hospital, Harvard Medical School, Boston MA, USA; Department of Operations Research and Financial Engineering, Princeton University, Princeton, NJ; Department of Experimental Cardiology, Heart Center, Amsterdam Cardiovascular Sciences, Amsterdam UMC location University of Amsterdam, Amsterdam, Netherlands; Institute for Molecular Medicine Finland (FIMM), Helsinki Institute of Life Science (HiLIFE), University of Helsinki, Helsinki, Finland; Division of Cardiology, Department of Medicine, Emory University School of Medicine, Atlanta, GA; Cardiology Division, Montefiore Healthcare Network/Albert Einstein College of Medicine, Bronx, NY; Cardiovascular Institute, Perelman School of Medicine, University of Pennsylvania, Philadelphia, PA; Department of Medicine, Division of Cardiology, Ted Rogers Program in Cardiotoxicity Prevention, Peter Munk Cardiac Centre, Toronto General Hospital, University Health Network, University of Toronto, Toronto, Ontario, Canada; Cardiology Division, Department of Medicine, Massachusetts General Hospital, Boston, MA; Department of Medicine, Massachusetts General Hospital, Harvard Medical School, Boston, MA; Cardiovascular Imaging Research Center (CIRC), Department of Radiology and Division of Cardiology, Massachusetts General Hospital, Boston, MA; Division of Cardiovascular Medicine, Department of Medicine, Brigham and Women’s Hospital, Boston, MA; Department of Medicine, Harvard Medical School, Boston, MA; Verve Therapeutics, Boston, MA; Demoulas Center for Cardiac Arrhythmias, Massachusetts General Hospital, Boston MA, USA

**Author notes:** Contributed equally to this work (Dimitri J. Maamari, Kiran J. Biddinger, Sean J. Jurgens). Jointly supervised this work (Amit V. Khera, Patrick T. Ellinor, Krishna G. Aragam). **Correspondence:** Krishna G. Aragam, MD, MS Massachusetts General Hospital Simches Research Building, 3.128 185 Cambridge Street Boston, MA 02114.

## Abstract

**Background:** Rare (monogenic) variants linked to non-ischemic dilated cardiomyopathy (DCM) are enriched among individuals with peripartum (PPCM), alcohol-induced (ALCM), and cancer therapy-related (CCM) cardiomyopathies, but are present in less than 15% of cases. Whether a common variant (polygenic) predisposition to DCM also pervades these secondary cardiomyopathies remains unclear.

**Methods:** We evaluated the association of a DCM polygenic score with PPCM, ALCM, and CCM in the Mass General Brigham (MGB) Biobank (n = 42,137), with replication in the UK Biobank (n = 295,160) and FinnGen (n = 417,950). We then assessed the proportion of cases with a monogenic variant and/or a high polygenic score (defined as > 80^th^ percentile of the score distribution). Finally, we queried medical charts to ascertain whether cardiomyopathy onset in those at high polygenic risk might have been heralded by relevant clinical risk factors.

**Results:** We identified 415 individuals with a secondary cardiomyopathy (30 with PPCM, 275 with ALCM, and 110 for CCM) across the three cohorts. The DCM polygenic score associated with PPCM (OR = 1.88 per 1 standard deviation (SD) increase in polygenic score, p= 0.001), ALCM (OR per SD = 1.38, p = 1.46E-07), and CCM (OR per SD = 1.58, p = 2.97E-06). Monogenic DCM variants were strongly associated with PPCM, ALCM, and CCM, but were present in less than 10% of cases. Roughly 40% of all secondary cardiomyopathy cases had a high polygenic score, which conferred ∼3-fold odds of cardiomyopathy (p <0.001). Most secondary cardiomyopathy cases lacked known antecedent clinical risk factors.

**Conclusion:** Cases of PPCM, ALCM, and CCM are enriched for monogenic DCM variants and a high DCM polygenic score, further supporting a shared genetic susceptibility influenced by distinct environmental precipitants. Considering both monogenic and polygenic risk for DCM may improve identification of individuals predisposed to secondary cardiomyopathies, particularly among those lacking established clinical risk factors.

## INTRODUCTION

Cardiomyopathies secondary to the peripartum state (PPCM), alcohol use (ALCM), and cancer therapy (CCM) are distinct clinical entities arising from specific physiological or environmental stressors. Each condition bears a significant clinical burden and confers a morbid prognosis.(1–3) However, recent studies have suggested that the genetic underpinnings of PPCM, ALCM, and CCM resemble those of non-ischemic dilated cardiomyopathy (DCM).(4–11) Specifically, rare, monogenic variants associated with DCM, such as truncating variants in *TTN*, have been found more frequently in secondary cardiomyopathy cases than in the broader population. These observations have supported a “two-hit hypothesis” wherein a genetic predisposition for cardiomyopathy combines with a subsequent stressor, such as pregnancy, alcohol, or chemotherapy, to transition a latent susceptibility to overt clinical disease. In addition, the findings have alluded to a role for genetic markers in the prognostication of PPCM, ALCM, and CCM, conditions for which our current clinical tools to assess upstream risk are lacking.

Despite being more prevalent in individuals with cardiomyopathies than in those without, rare monogenic DCM variants are present in less than 15% of PPCM, ALCM, and CCM cases.(4–11) In recent years, common genetic variants have been shown to contribute substantially to heritable risk for complex diseases such as DCM.(12–20) In fact, a DCM common-variant (polygenic) score appears to influence DCM risk in both carriers and non-carriers of relevant monogenic variants.(21) Whether this polygenic liability to DCM also contributes to risk of PPCM, ALCM, and CCM remains to be explored.

In this study, we examined whether a polygenic predisposition to DCM underlies PPCM, ALCM, and CCM. First, through medical chart review in a health system-associated biobank, we identified cases of secondary cardiomyopathies and assessed for the presence of antecedent clinical risk factors. Next, we tested the link between a DCM polygenic score and PPCM, ALCM, and CCM across three independent biobank populations. We then identified secondary cardiomyopathy cases harboring rare, monogenic DCM variants, and examined for overlap with polygenic risk. Finally, we re-examined the charts of cases at high polygenic risk to determine whether any aspects of their clinical history or presentation might have foretold their elevated risk of disease.

## METHODS

### Study subjects

We studied participants from the Mass General Brigham (MGB) Biobank, the UK Biobank, and FinnGen. The MGB Biobank is a large biospecimen and data repository based in Eastern Massachusetts, largely comprising patients from the Massachusetts General Hospital and the Brigham and Women’s Hospital.(22–24) Genomic data were available for 50,623 participants after quality control at the time of this analysis, with both array-based genotyping and whole-exome sequencing data. The final analyses were performed on 42,435 MGB Biobank participants, as detailed below. The UK Biobank is a prospective national biobank study with genetic and phenotypic data collected between 2006 and 2010 on approximately 500,000 individuals aged 40-69 at recruitment.(25) A total of 438,069 individuals were included for final analysis after quality control. FinnGen is a nationwide collection of prospective Finnish epidemiological and disease-based cohorts and hospital biobank samples.(26) Data from 453,733 participants from FinnGen Data Freeze 11 were used for this study. A complete description of the participating studies can be found in the **Supplementary Methods**

Informed consent was collected from all participants. Analyses were performed under IRB protocol 2018P002292 for MGB Biobank and application number 17488 for UK Biobank.

### Curation of cardiomyopathy phenotypes

In FinnGen and UK Biobank, secondary cardiomyopathy cases were defined based on the presence or absence of relevant International Classification of Diseases, Tenth Revision (ICD-10) billing codes: O90.3 (“peripartum cardiomyopathy”) for PPCM, I42.6 (“alcoholic cardiomyopathy”) for ALCM, and I42.7 (“cardiomyopathy due to drugs and other external agents”) as a surrogate for CCM. In MGB Biobank, we imposed more stringent phenotypic criteria given access to the medical records of all study participants. Specifically, we first nominated secondary cardiomyopathy cases using the above ICD-10 codes; for CCM, we applied a modified criteria consisting of ICD-10 codes I42.7 (“cardiomyopathy due to drugs and other external agents”), I42.0 (“dilated cardiomyopathy”) or I50.1 (“left ventricular failure”) and any history of antecedent anthracycline use as indicated by an automated biobank classifier. We then pursued chart validation by two medical doctors blinded to the genetic data for PPCM (D.J.M., A.A.S.), ALCM (D.J.M., D.H.), and CCM (D.J.M., C.A.G) to confirm each nominated case. We also defined DCM in MGB Biobank using ICD-10 code I42.0 (“dilated cardiomyopathy”) and excluding individuals with antecedent ischemic heart disease. The control group comprised participants without heart failure (ICD-10 code I50) or cardiomyopathy (ICD-10 code I42).

### Genotyping arrays and polygenic score calculation

Array data in the MGB Biobank were available via the Illumina Global Screening Array. Array data in the UK Biobank were available using the Affymetrix UK BiLEVE Axiom array and the Affymetrix UK Biobank Axiom® array. Array data in FinnGen were generated using Illumina and Affymetrix arrays (Illumina Inc., San Diego, and Thermo Fisher Scientific, Santa Clara, CA, USA). Genotype imputation in FinnGen was performed using a population-specific SISu v4 imputation reference panel of 8,557 whole genomes as detailed in a public protocol (https://www.protocols.io/view/genotype-imputation-workflow-v3-0-e6nvw78dlmkj/v2).

We previously conducted genome-wide association studies of cardiac magnetic resonance imaging-derived measures of left ventricular size and function, quantitative endophenotypes for DCM.(15) These data have been leveraged to generate genome-wide polygenic scores (GPS) for DCM using the LDPred2 computational algorithm.(27) Here, we employed one of the developed and validated GPS for DCM (using LV end-systolic volume indexed for body surface area; LVESVi), comprising 1,137,406 common genetic variants, each with a minor allele frequency greater than 1% (**Supplementary Methods**).(15,28)

### Analyses of rare monogenic variants

Analyses of rare, monogenic variants of relevance to DCM were conducted from the MGB Biobank with whole exome sequencing data as previously described.(29,30) Briefly, we prioritized ClinVar pathogenic or likely pathogenic (P/LP) variants from 12 genes with “definitive” or “strong” evidence for DCM pathogenicity, as determined by the international ClinGen consortium: *BAG3*, *DES*, *DSP*, *FLNC*, *LMNA*, *MYH7*, *PLN*, *RBM20*, *SCN5A*, *TNNC1*, *TNNT2*, and *TTN.* For 6 of the 12 genes mentioned, we also included loss-of-function (LOF) variants based on evidence supporting protein truncation as a mechanism for DCM: *BAG3,*(31,32) *DSP,*(33) *FLNC,*(34) *LMNA,*(35) *RBM20,*(36) and *TTN.*(35,37) For *TTN*, LOF variants were restricted to those affecting cardiac isoforms.(29) In a sensitivity analysis, we evaluated rare variants in an expanded set of 91 genes included on clinical gene-screening panels for DCM to assess whether this might significantly increase the estimated contribution of rare monogenic variants to secondary cardiomyopathy risk. (**Supplementary Methods**)

### Analysis of combined genetic and clinical risk

Among individuals with chart-validated secondary cardiomyopathies in MGB Biobank, we explored clinical presentations and antecedent risk factors through detailed chart reviews (**Supplementary Tables 10, 11 and 12**). For all confirmed cases of PPCM, ALCM, and CCM in the MGB Biobank, we used a combination of ICD-10 codes and medical record chart review to obtain individuals’ medical history, gender, age at the time of the analysis, race, ethnicity, presence or absence of hypertension, diabetes mellitus or obesity at the time of diagnosis. A family history of cardiomyopathy was ascertained through review of clinician notes and other relevant documentation in the electronic medical record. We also obtained clinical information relevant to each specific cardiomyopathy; i.e. for PPCM, the presence of preeclampsia or eclampsia, and for CCM, the type of cancer and anthracycline agent used.

### Statistical analysis

Statistical analysis was carried out using R software version 4.1.0 and PLINK 2.0. Statistics were presented as proportions for categorical variables and as means with standard deviations or medians with interquartile range for continuous variables. A chi-square test of independence was used for categorical variables. An unpaired t-test was used for continuous variables, with the level of statistical significance set at P <0.05. Associations of relevant monogenic variants and the polygenic score with secondary cardiomyopathies were evaluated via logistic regression adjusting for age, sex, genotyping array, and the top ten principal components of genetic ancestry.

## RESULTS

### Phenotype validation and baseline characteristics in MGB Biobank

Among secondary cardiomyopathy cases in MGB Biobank identified by electronic phenotyping, 21 of 26 (80.8%) PPCM cases, 76 out of 101 (75.2%) ALCM cases, and 16 of 57 (28.1%) CCM cases were confirmed by chart review. The most common reasons for misclassification were the diagnosis of another cardiomyopathy (e.g. ischemic, hypertrophic, or tachycardia-induced cardiomyopathies) concluded to be primary by chart review, erroneous ICD-10 coding, or ambiguous temporality between cardiomyopathy diagnosis and receipt of anthracycline therapy.

Baseline characteristics of chart-confirmed cases of PPCM, ALCM, and CCM as compared to controls in the MGB Biobank are presented in **Table 1**. All individuals with PPCM were females, and their mean age was 46 years at the time of the analysis. Individuals with ALCM had the highest proportion of males at 82.9%, and the highest mean age at 65.3 years. Half of the individuals with CCM were females; the mean age was 58.6 years. About half of the individuals with PPCM (47.6%) and CCM (56.2%) identified as being White, with 28.6% of those with PPCM and 25% of those with CCM identifying as being Black or African American. The vast majority of those with ALCM (81.6%) identified as being White.

**Table 1:** Baseline characteristics of the MGB Biobank study population. ALCM = alcohol-induced cardiomyopathy; CCM = cancer therapy-induced cardiomyopathy; F = female; PPCM = peripartum cardiomyopathy; SD = standard deviation

|  | **PPCM**<br>**(n=21)** | **ALCM**<br>**(n=76)** | **CCM**<br>**(n=16)** | **Control**<br>**(n=42,024)** | **P-value** |
| --- | --- | --- | --- | --- | --- |
| **Gender = F (%)** | 21 (100.0) | 13 (17.1) | 8 (50.0) | 24,509 (58.3) | <0.001 |
| **Mean age (SD)** | 46.4 (9.2) | 65.3 (11.3) | 58.5 (19.3) | 55.9 (17.0) | <0.001 |
| **Race (%)** |  |  |  |  | <0.001 |
| **Asian** | 0 (0.0) | 1 (1.3) | 2 (12.5) | 1,311 (3.1) |  |
| **Black or**<br>**African**<br>**American** | 6 (28.6) | 8 (10.5) | 4 (25.0) | 1,920 (4.6) |  |
| **White** | 10 (47.6) | 62 (81.6) | 9 (56.2) | 35,123 (83.6) |  |
| **Other** | 5 (23.8) | 5 (6.6) | 1 (6.2) | 3,670 (8.7) |  |
| **Ethnicity (%)** |  |  |  |  | 0.029 |
| **Hispanic** | 2 (9.5) | 6 (7.9) | 0 (0.0) | 1,314 (3.1) |  |
| **Non-**<br>**Hispanic or**<br>**unknown** | 19 (90.5) | 70 (92.1) | 16 (100.0) | 40,710 (96.9) |  |

### Clinical risk factors for secondary cardiomyopathy phenotypes

We assessed for the presence of clinical risk factors for cardiomyopathy at the time of diagnosis among individuals with secondary cardiomyopathy (**Figure 1, Supplementary Table 3**). Among those with PPCM, 5 (23.8%) had preeclampsia, 4 (19.0%) had hypertension, 4 (19.0%) had diabetes mellitus and 2 (9.5%) were obese. None had a family history of cardiomyopathy. For ALCM, 47 (61.8%) had hypertension, 20 (26.3%) had diabetes mellitus, 9 (11.8%) were obese, and 5 (6.6%) had a family history of cardiomyopathy at the time of diagnosis. For CCM, 2 (12.5%) had hypertension, 2 (12.5%) had diabetes mellitus, 2 (12.5%) had a family history of cardiomyopathy and none were obese at the time of diagnosis. Overall, 57.1%, 28.9%, and 81.2% of those with PPCM, ALCM, and CCM, respectively, had no diagnosed clinical risk factor at the time of diagnosis of their secondary cardiomyopathy (**Figure 1**). Only 4.8% of those with PPCM, 2.6% with ALCM, and 6.2% with CCM had three clinical risk factors at the time of diagnosis of their cardiomyopathies.

**Figure 1:**
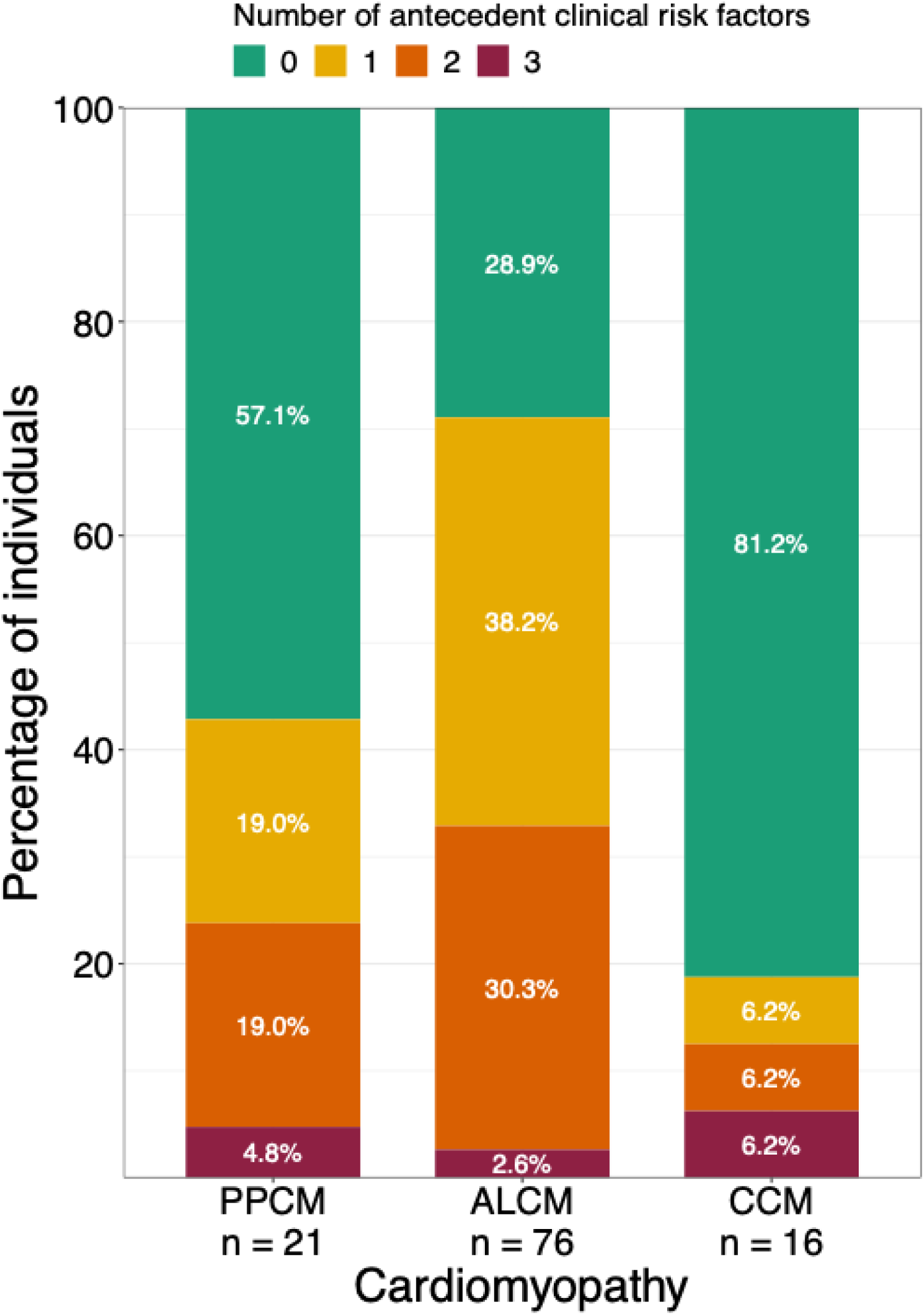
Distribution of participants by number of antecedent clinical risk factors for cardiomyopathy. This figure displays the number of participants with 0, 1, 2, or 3 clinical risk factors present at the time of secondary cardiomyopathy diagnosis. Risk factors include hypertension, diabetes mellitus, obesity, or a documented family history of DCM. For participants with PPCM, preeclampsia was also included as a risk factor. ALCM = alcohol-induced cardiomyopathy; CCM = cancer therapy-induced cardiomyopathy; PPCM = peripartum cardiomyopathy.

### Polygenic score associations with secondary cardiomyopathies

In MGB Biobank, the DCM GPS associated with chart-validated cases of PPCM (OR per SD = 1.78, P = 0.013), ALCM (OR per SD = 1.99, P = 9.46E-09), and CCM (OR per SD = 1.78, P = 0.03) (**Figure 2, Supplementary Table 4**). The GPS also demonstrated associations or trends with secondary cardiomyopathy phenotypes in the MGB cohort without chart-validation, though to a lesser extent (PPCM: OR per SD = 1.55, P = 0.035; ALCM: OR per SD = 1.63, P = 2.06E-06; CCM: OR per SD = 1.20, P = 0.180). We sought replication in the UK Biobank and FinnGen, which included 302 cases of secondary cardiomyopathies ascertained by electronic phenotyping (see **Supplementary Tables 1** and **2** for baseline characteristics in UK Biobank and FinnGen). In both UK Biobank and FinnGen, GPS associations were consistent with those in MGB Biobank for all three phenotypes (**Figure 2**; **Supplementary Tables 5** and **6**). A meta-analysis across biobanks – including a total of 415 secondary cardiomyopathy cases and 754,832 controls – demonstrated strong associations between a higher polygenic score and increased risk of PPCM (OR per SD = 1.89, P = 0.001, n = 30), ALCM (OR per SD = 1.38, P = 1.46E-07, n = 275), and CCM (OR per SD = 1.58, P = 2.97E-06, n = 110) (**Figure 2**).

**Figure 2:**
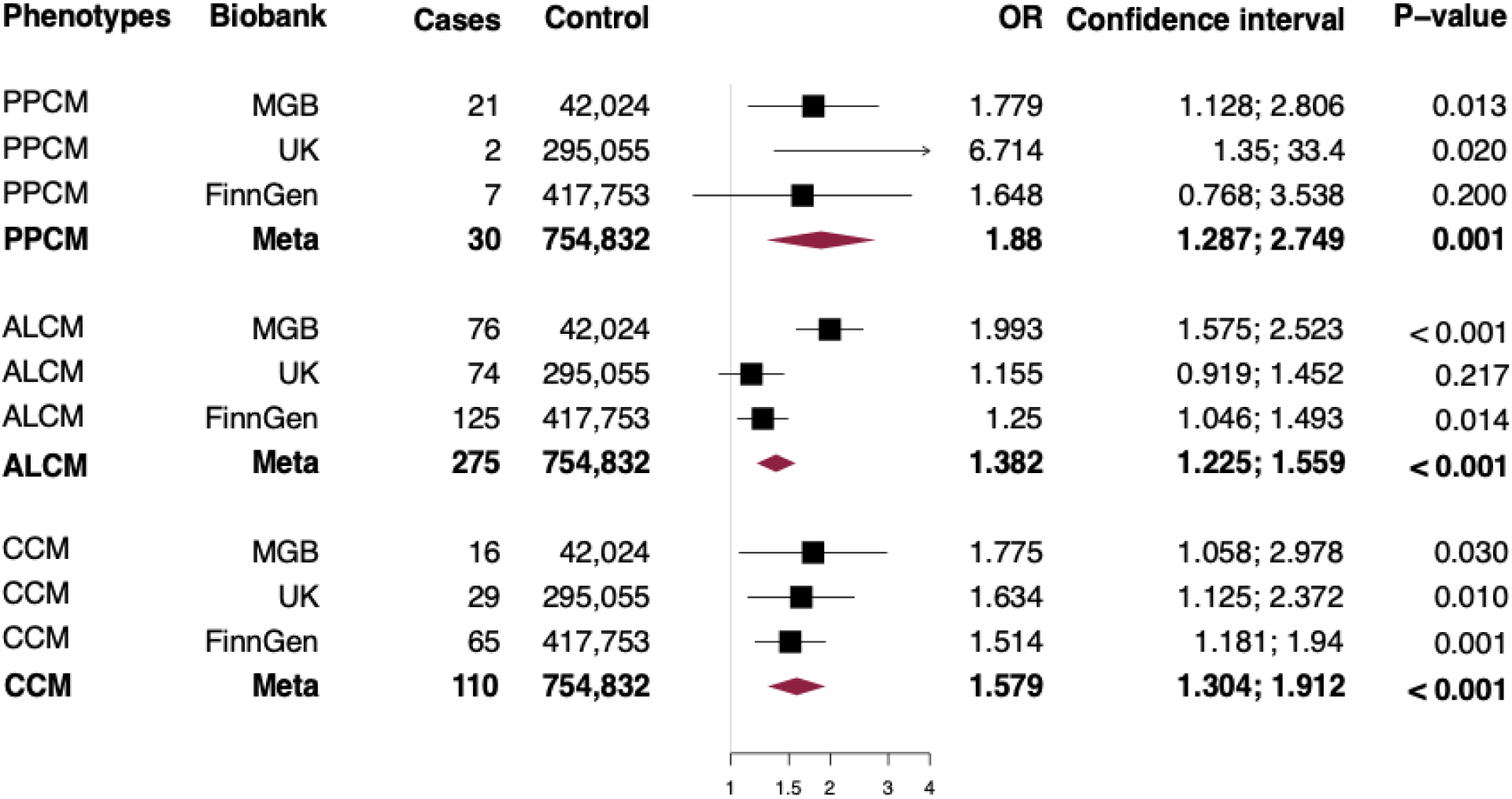
Association of DCM polygenic score with PPCM, ALCM, and CCM. Forest plots depict the odds of developing the specified cardiomyopathy per 1 standard deviation increase in the polygenic score across the MGB Biobank, UK Biobank, FinnGen, and a meta-analysis of all three cohorts. Effect estimates were derived from logistic regression models adjusted for age, sex, and the first ten principal components. ALCM = alcoholic cardiomyopathy; CCM = cancer chemotherapy-induced cardiomyopathy; PPCM = peripartum cardiomyopathy.

We focused thereafter on chart-validated cases in MGB Biobank, and found that the distribution of the DCM GPS was similar for PPCM (median GPS percentile among study population = 74), ALCM (median GPS percentile = 72), and CCM (median GPS percentile = 73). Of note, GPS distributions for secondary cardiomyopathy cases resembled that of DCM (total N = 298; median GPS percentile = 67), with all cardiomyopathies enriched for a high polygenic score as compared to controls (median GPS percentile = 50) (**Figure 3**).

**Figure 3:**
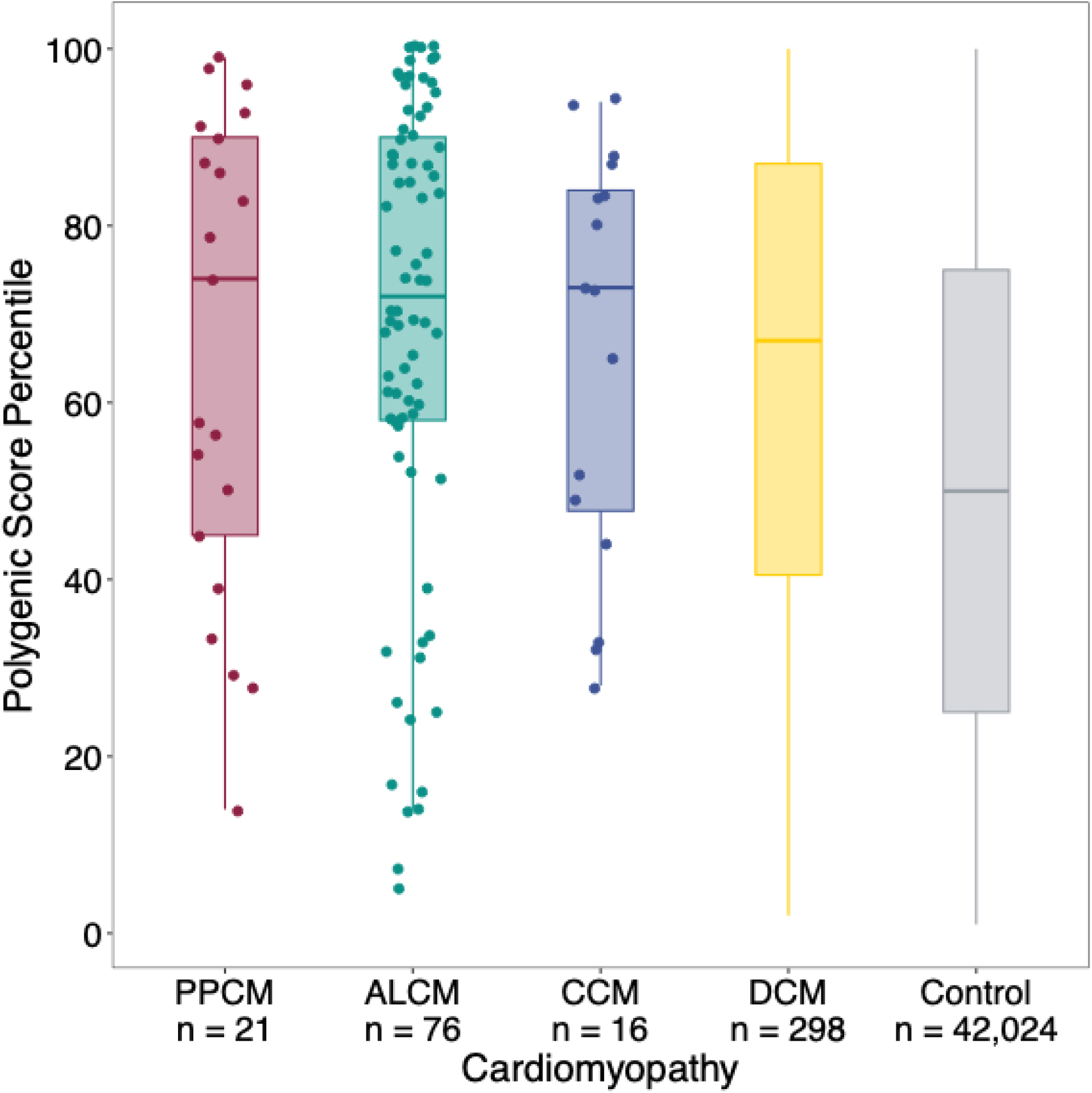
Distribution of DCM polygenic score in individuals with a secondary cardiomyopathy or DCM in the MGB Biobank. Boxplots illustrate the distribution of a DCM polygenic score among individuals with PPCM, ALCM, CCM, DCM and a control cohort in the MGB Biobank. PPCM = peripartum cardiomyopathy; ALCM = alcoholic cardiomyopathy; CCM = cancer chemotherapy-induced cardiomyopathy; DCM = dilated cardiomyopathy.

In MGB Biobank, we further assessed the risk of secondary cardiomyopathies associated with being in an upper stratum of the DCM GPS, using two different cutoffs: top 5^th^ percentile and top 20^th^ percentile of the GPS distribution. Membership in the top strata of each cutoff was associated with a significant increased risk of cardiomyopathy in the combined secondary cardiomyopathy cohort (top 5^th^ percentile OR =3.34, P <0.001, top 20^th^ percentile OR = 2.98, P <0.001). For each individual cardiomyopathy, membership in the top 20% of the DCM GPS was associated with higher odds of PPCM (OR = 3.15, P = 0.01) and ALCM (OR = 2.98, P = 2.99E-06) when compared to those with a polygenic score at or below the 80^th^ percentile; a similar trend was seen for CCM (OR = 2.61, P = 0.065) (**Supplementary Table 7**). Since membership in the top 20^th^ percentile of the DCM GPS offered sufficient power and revealed similar (roughly 3-fold) odds of disease across each of the three secondary cardiomyopathies, this threshold was used as an exemplar of “high polygenic risk.” As expected, secondary cardiomyopathy cases were enriched for a high polygenic score, which was present in 9 (42.9%) individuals with PPCM, 32 (42.1%) with ALCM, and 6 (37.5%) of those with CCM.

### Monogenic variant associations in MGB Biobank for chart-validated phenotypes

Next, we examined the prevalence and associated risk of DCM monogenic variants among 42,137 individuals (113 cases and 42,024 controls) in MGB Biobank. Of the 12 genes assessed, we found only carriers of high-confidence *TTN* truncating variants (*TTN*tv) among secondary cardiomyopathy cases. Specifically, we observed *TTN*tv in 2 PPCM cases (9.5%), 4 ALCM cases (5.3%), and one CCM case (6.3%). Despite the small sample size of cases, we recapitulated strong associations between DCM monogenic mutation carriers and PPCM (OR = 27.22, P = 2.79E-05, 95% CI 5.80-127.64), ALCM (OR = 9.5, P = 1.87E-05, 95% CI 3.39-26.67) and CCM (OR = 13.9, P = 0.012, 95% CI 1.79-108.60) (**Supplementary Table 8**). In a sensitivity analysis considering an expanded list of 91 genes of potential relevance to DCM, only one additional secondary cardiomyopathy case harbored a relevant monogenic variant (ALCM-28 with a *FKTN* variant).

### Combined polygenic and monogenic assessment for chart-validated phenotypes

We evaluated the proportion of chart-validated secondary cardiomyopathy cases in MGB Biobank with a monogenic variant, a high polygenic score (defined as being in the top 20% of the GPS distribution with ∼3-fold disease risk as seen above), or both (**Figure 4, Supplementary Table 9**). Among individuals with PPCM, 38.1% had a high GPS only, 4.8% harbored a monogenic DCM variant only, and 4.8% had both a high GPS and a monogenic variant. Of those with ALCM, 39.5% had a high GPS only, 2.6% harbored a monogenic variant only, and 2.6% had both a monogenic variant and a high GPS. Among those with CCM, 31.3% had a high GPS only, no cases harbored a monogenic variant only, and 6.3% had both a monogenic DCM variant and a high GPS.

**Figure 4:**
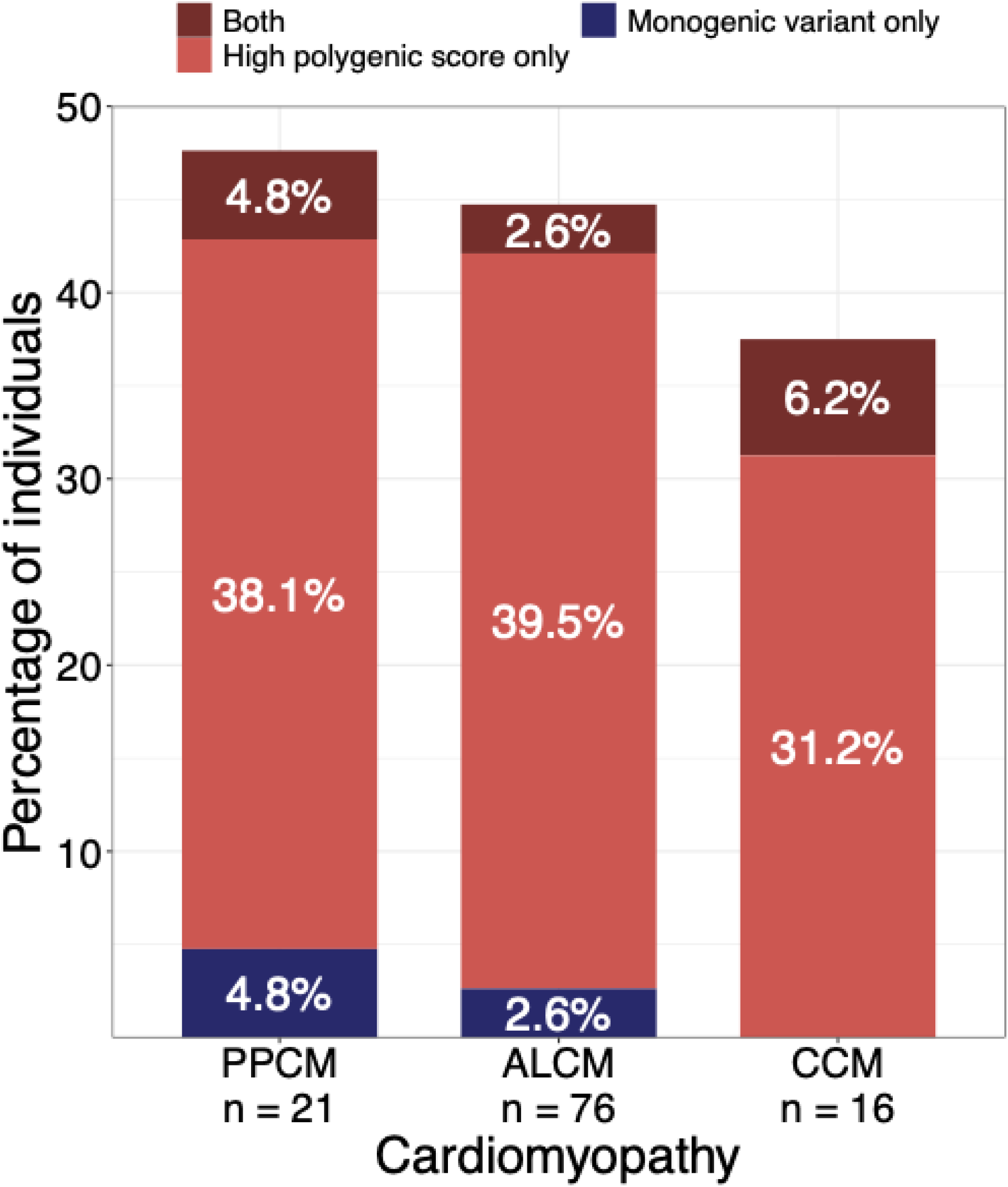
Relative contributions of monogenic and polygenic risk in secondary cardiomyopathies. The figure presents the number and percentage of individuals categorized into three classifications of genetic risk: those with a high polygenic score only (defined as a score ≥80^th^ percentile), those with a DCM monogenic variant only, and those with both a high polygenic score and a monogenic variant.

### Clinical histories of cases with high polygenic risk

We reviewed the histories of secondary cardiomyopathy cases found to have high polygenic risk to explore their clinical presentations and whether any antecedent clinical factors might have foretold their conditions (**Supplementary Table 3**). A high polygenic score was defined as being at or above the 80^th^ percentile of the GPS distribution (**Supplementary Table 7**). Of the 9 PPCM cases with a high polygenic score, 2 had a prior history of hypertension and concomitant obesity, and 2 developed pre-eclampsia during pregnancy; the majority lacked any identifiable clinical risk factor for cardiomyopathy onset (**Table 2, Supplementary Table 10**). Of the 32 ALCM cases with a high polygenic score, a large proportion had co-morbid risk factors like diabetes or hypertension, consistent with the older mean age of this population (**Table 3, Supplementary Table 11**). However, these risk factors were typically chronic, treated, and not thought to be the primary etiology of cardiomyopathy. Instead, a presumptive diagnosis of ALCM was made for many cases (ALCM-2, ALCM-3, ALCM-5, and ALCM-14). One individual with ALCM (ALCM-22) harbored a *TTNtv* and a high polygenic score – this individual manifested early-onset systolic dysfunction in their 20’s in the setting of heavy alcohol consumption and went on to require heart transplantation. Notably, three ALCM cases with a high polygenic score had a documented family history of cardiomyopathy. In fact, two of those individuals were seen by cardiologists, who hypothesized that the cardiomyopathy was due to a combination of heavy alcohol use and a “familial” predisposition (ALCM-9, ALCM-27). However, none of these ALCM cases harbored an identifiable monogenic variant for DCM in our analysis and therefore would likely not have been identified as having a genetic etiology by clinical genetic tests currently available for DCM. Of the 6 individuals with CCM and a high polygenic score, none had a relevant antecedent clinical risk factor (**Table 4, Supplementary Table 12**).

**Table 2:** Brief clinical histories and monogenic variant status for individuals with peripartum cardiomyopathy and a polygenic score greater than or equal to the 80^th^ percentile. DCM = dilated cardiomyopathy; Dx = diagnosed; F = female; IVF = in-vitro fertilization; PPCM = peripartum cardiomyopathy; RV = right ventricle; VUS = variant of uncertain significance.

| Case Number | Gender | Monogenic variant | Polygenic score percentile | Clinical information | Preeclampsia | Hypertension | Diabetes mellitus | Obesity | Family history of DCM |
| --- | --- | --- | --- | --- | --- | --- | --- | --- | --- |
| PPCM-1 | F | No | 99 | Dx in their 50's after IVF | Yes | No | No | No | No |
| PPCM-2 | F | No | 98 | Cardiac stress test without evidence of ischemia | No | Yes | No | Yes | No |
| PPCM-3 | F | No | 96 | Dx in their 20's | No | Yes | No | Yes | No |
| PPCM-4 | F | No | 93 | Dx in their 30's | No | No | No | No | No |
| PPCM-5 | F | No | 91 | Dx in their 20's, RV biopsy without evidence of active myocarditis | No | No | No | No | No |
| PPCM-6 | F | No | 90 | Dx in their 20's, *TTN* VUS | No | No | No | No | No |
| PPCM-7 | F | Yes, *TTN* LOF | 87 | Dx in their 20's | No | No | No | No | No |
| PPCM-8 | F | No | 86 | Dx in their 30's | Yes | No | No | No | No |
| PPCM-9 | F | No | 83 | Dx in their 40's | No | No | No | No | No |

**Table 3:** Brief clinical histories and monogenic variant status for individuals with alcohol-induced cardiomyopathy and a polygenic score greater than or equal to the 80^th^ percentile. ALCM = alcohol-induced cardiomyopathy; AF = atrial fibrillation; DCM = dilated cardiomyopathy; F = female; HF = heart failure; LVEF = left ventricular ejection fraction; M = male; MRI = magnetic resonance imaging.

| Case number | Gender | Monogenic variant | Polygenic score percentile | Clinical information | Hypertension | Diabetes mellitus | Obesity | Family history of DCM |
| --- | --- | --- | --- | --- | --- | --- | --- | --- |
| ALCM-1 | M | No | 100 | LVEF 15% | Yes | Yes | No | No |
| ALCM-2 | M | No | 100 | LVEF 19% | No | No | No | No |
| ALCM-3 | M | No | 100 | LVEF 15% | Yes | Yes | No | No |
| ALCM-4 | M | No | 100 | LVEF 15% | Yes | Yes | No | No |
| ALCM-5 | M | Yes, *TTN* LOF | 99 | Biventricular HF, LVEF 26%, coronary angiogram with only luminal irregularities | Yes | No | No | No |
| ALCM-6 | M | No | 99 | LVEF 35%, 4 glasses of wine daily | No | No | No | No |
| ALCM-7 | M | No | 99 | Biventricular HF, LVEF 25% | Yes | Yes | No | No |
| ALCM-8 | F | No | 97 | LVEF 16% | Yes | No | No | No |
| ALCM-9 | M | No | 97 | LVEF 9%, four chamber dilation, clinical suspicion for familial DCM | No | No | No | Yes |
| ALCM-10 | M | No | 97 | LVEF 26%, diffusely hypokinetic right and left ventricles | Yes | No | No | No |
| ALCM-11 | M | No | 97 | LVEF 44% | No | No | No | No |
| ALCM-12 | F | No | 96 | LVEF 50% | No | No | Yes | No |
| ALCM-13 | M | No | 95 | LVEF 10% | Yes | No | No | No |
| ALCM-14 | M | No | 94 | LVEF 21%, more than 12 drinks daily at the time of diagnosis | Yes | Yes | No | No |
| ALCM-15 | M | No | 93 | LVEF 10% | No | No | No | No |
| ALCM-16 | M | No | 93 | LVEF 22% | No | No | No | No |
| ALCM-17 | M | No | 92 | LVEF 26% | Yes | No | No | No |
| ALCM-18 | M | No | 91 | LVEF 40% | Yes | No | Yes | No |
| ALCM-19 | M | No | 90 | LVEF 33%, 5 drinks of hard liquor daily at the time of diagnosis; father had DCM | No | No | No | Yes |
| ALCM-20 | M | No | 90 | LVEF 45% | Yes | No | No | No |
| ALCM-21 | M | No | 89 | LVEF 15% | Yes | No | No | No |
| ALCM-22 | M | Yes, *TTN*<br>LOF | 88 | HF in their 20's, required heart transplantation | No | No | No | No |
| ALCM-23 | M | No | 88 | LVEF 20% | No | Yes | No | No |
| ALCM-24 | M | No | 87 | HF in their 40's, exercise myocardial perfusion imaging without evidence of ischemia | Yes | No | No | No |
| ALCM-25 | M | No | 87 | LVEF 20% | Yes | No | No | No |
| ALCM-26 | F | No | 87 | LVEF 48% by cardiac MRI | Yes | No | No | No |
| ALCM-27 | M | No | 86 | LVEF 23%, HF specialist thought HF secondary to heavy alcohol use and possible genetic predisposition due to family history of DCM | Yes | No | No | Yes |
| ALCM-28 | M | No | 85 | LVEF 16%, ALCM as supported by diffuse hypokinesis and alcohol use | Yes | Yes | No | No |
| ALCM-29 | M | No | 85 | LVEF 34%, ALCM as supported by global hypokinesis and alcohol use | No | No | No | No |
| ALCM-30 | M | No | 84 | LVEF 15% | Yes | Yes | No | No |
| ALCM-31 | M | No | 82 | LVEF 30%, ischemic evaluation unrevealing | Yes | No | No | No |
| ALCM-32 | F | No | 82 | LVEF 30%, alcohol-induced cirrhosis | No | No | No | No |

**Table 4:** Brief clinical histories and monogenic variant status for individuals with cancer therapy-induced cardiomyopathy and a polygenic score greater than or equal to the 80^th^ percentile. CCM = cancer therapy-induced cardiomyopathy; DCM = dilated cardiomyopathy; F = female; HF = heart failure; LVEF = left ventricular ejection fraction; M = male.

| Case number | Monogenic variant | Polygenic score percentile | Clinical information | Cardiotoxic chemo-therapeutic agent(s) | Malignancy | Hypertension | Diabetes mellitus | Obesity | Family history of DCM |
| --- | --- | --- | --- | --- | --- | --- | --- | --- | --- |
| CCM-1 | No | 94 | LVEF 66% to 50% after the fourth cycle of anthracyclines | Anthracycline and trastuzumab | Invasive ductal carcinoma of the breast | No | No | No | No |
| CCM-2 | Yes, *TTN* LOF | 94 | HF 15 years after completion of 4 cycles of anthracyclines and radiation to the left breast, ischemic and other non-ischemic evaluations unrevealing | Doxorubicin | Ductal carcinoma of the breast | No | No | No | No |
| CCM-3 | No | 88 | LVEF 45-50% after completion of chemotherapy | Doxorubicin and trastuzumab | Breast cancer | No | No | No | No |
| CCM-4 | No | 87 | LVEF 32% 10 years after treatment with anthracyclines | Doxorubicin | Breast cancer | No | No | No | No |
| CCM-5 | No | 83 | LVEF 49% 10 years after completing a treatment with anthracyclines | Doxorubicin | Osteosarcoma | No | No | No | No |
| CCM-6 | No | 83 | LVEF 46% after treatment with anthracyclines | Doxorubicin | Osteosarcoma | No | No | No | No |

## DISCUSSION

In this study across three distinct cohorts, we found that individuals with PPCM, ALCM, and CCM were enriched for a high polygenic predisposition to DCM. This polygenic liability was more commonly observed among cases than was the presence of a monogenic DCM variant (i.e. *TTNtv*). Moreover, a high polygenic score was often the only identifiable risk factor for cardiomyopathy, as most cases did not present with other established clinical risk factors prior to disease onset.

Studies have shown that rare genetic variants linked to DCM, such as *TTNtv*, are enriched among cases of PPCM, ALCM, and CCM compared to the general population, contributing to 7% to 15% of cases.(4,8,9,11) We now demonstrate a shared common-variant (polygenic) signature that further supports a unifying genetic basis across these cardiomyopathies. A recent systematic review and meta-analysis of pregnant cancer survivors found that a history of cancer therapy-related cardiac dysfunction was associated with 47-fold higher odds of subsequent peripartum left ventricular dysfunction or heart failure.(38) A genetic similarity may contribute to the striking overlap in risk between these seemingly disparate cardiomyopathies. Taken together, a history of one cardiomyopathy may indicate an elevated risk for another, potentially justifying longitudinal surveillance for distinct cardiomyopathies in individuals who have recovered from an initial event.

Familial screening is recommended for DCM probands due to the substantially elevated risk of DCM among first-degree relatives.(39) The enrichment of rare pathogenic mutations in these relatives has been documented, affirming a genetic contribution to familial disease risk.(40,41) We recently demonstrated that polygenic factors may contribute more to disease risk among “genotype-negative” DCM cases (i.e. those without a rare mutation).(19) Similarly, in this study, we observed cases of ALCM and CCM with a “family history of cardiomyopathy” that lacked a monogenic DCM variant but had an elevated DCM polygenic score. These findings suggest that familial screening should be considered not only for DCM probands but also for probands of these secondary cardiomyopathies. Additionally, they indicate that assessing a DCM polygenic score alongside traditional monogenic DCM variants could enhance the detection of genetic risk for PPCM, ALCM, and CCM.

The value of genetic information for disease prognostication depends on whether it enhances established clinical predictors and whether early risk detection leads to changes in management. Several polygenic scores – such as those for coronary artery disease – have faced scrutiny due to questions about their added predictive benefit beyond already robust clinical risk models.(42–44) In contrast, individuals at risk for PPCM, ALCM, and CCM are not easily identified in the clinical setting. These cardiomyopathies often develop without obvious risk factors, as demonstrated in the current study, where most cases – particularly PPCM and CCM – lacked clinical antecedents at the time of diagnosis. Consequently, clinical risk prediction models for these cardiomyopathies have shown poor discriminatory performance (i.e. for CCM), have not been externally validated (i.e. for PPCM), or are altogether lacking (i.e. for ALCM).(45–48) A role for polygenic prediction may therefore be more tractable for such conditions, given the potential to improve upon our current, suboptimal standards for disease prognostication.

Improved risk assessment may in turn influence specific management decisions for these secondary cardiomyopathies. For example, while cardiac imaging is not routinely performed for pregnant women, it could be offered to those with a genetic predisposition (whether monogenic or polygenic) for cardiomyopathy. In cancer patients scheduled to receive anthracycline therapy, genetic risk might guide the frequency and timing of surveillance cardiac imaging and could even affect the choice to pursue an alternative chemotherapeutic regimen with lower cardiotoxic potential, if available. Furthermore, the proactive use of guideline-directed medical therapies might be considered for individuals with a monogenic variant or a high DCM polygenic score to prevent the onset of secondary cardiomyopathy.(49) However, the utility of genetic risk assessments in these and other contexts must be validated through analyses of prospective cohorts before they can be adopted into clinical practice.

Our analysis also highlights the utility of large biobanks in identifying cases of PPCM, ALCM, and CCM. Traditionally, studies of rarer cardiomyopathies have relied on the dedicated recruitment of cases, such as in the seminal, multi-site Investigations of Pregnancy-Associated Cardiomyopathy (IPAC) registry for PPCM.(50) While these cohort-based recruitment efforts remain essential, growing biobanks offer another valuable source to expand case collections and advance research on these important but less common conditions.

Our study has several limitations. First, as above, the use of electronic health record-based phenotyping alone to identify secondary cardiomyopathy cases within the UK and FinnGen Biobanks may have inherent constraints; although it should be noted that the GPS tracked with phenotypic versions in MGB that relied solely on ICD-10 codes and were not chart validated. Second, our DCM polygenic score was developed using data from the UK Biobank, which predominantly comprises individuals of European genetic ancestry. As a result, the reliability of the DCM polygenic score in associating with PPCM, ALCM, and CCM in individuals of non-European genetic backgrounds may be reduced. Third, our relatively small study population may limit the generalizability of our findings, although it should be noted that the current study is similar or larger in size as compared to other studies on these rare cardiomyopathies. Fourth, while the assessment of antecedent clinical risk factors was based on a detailed review of medical charts, some risk factors may not have been fully ascertained or documented in a routine clinical setting. Fifth, the current study was a retrospective analysis focused on establishing the relative enrichment of the DCM polygenic score among secondary cardiomyopathy cases versus controls. As mentioned earlier, prospective evaluations of the DCM polygenic score in relation to incident secondary cardiomyopathies are necessary before clinical implementation. Finally, the designation of a “high polygenic score” based on any percentile cutpoint is inherently arbitrary. In this study, the 80^th^ percentile threshold was chosen empirically, as it corresponded to an approximately three-fold increased risk of cardiomyopathy. However, this threshold serves only as a proof of concept and does not fully reflect the proportion of cardiomyopathy risk attributable to polygenic susceptibility. Future research is needed to identify optimal polygenic score thresholds for clinical risk stratification and risk mitigation strategies.

## CONCLUSION

A polygenic score for DCM is strongly associated with PPCM, ALCM, and CCM, further suggesting that these secondary cardiomyopathies arise from a shared genetic predisposition acting in concert with unique external stressors to the myocardium.

## Data Availability

All data produced in the present study are available upon reasonable request to the authors

## Acknowledgements

We thank all participants of the MGB Biobank, UK Biobank, and FinnGen, whose contributions made this study possible. K.G.A. was supported by the National Institutes of Health (1K08HL153937) and the American Heart Association (862032). P.T.E. received funding from the National Institutes of Health (1R01HL092577, 1R01HL157635, 5R01HL139731), the American Heart Association (18SFRN34110082), and the European Union (MAESTRIA 965286). Z.A. received funding from the National Institutes of Health (R01 HL152446). P.T. is supported by a Canada Research Chair in Cardiooncology (CRC-2019-00097) and the Canadian Cancer Society/Canadian Institutes of Health Research’s W. David Hargraft Grant. J.T.R. received support from a Sigrid Jusélius Fellowship. S.J.J. was supported by an Amsterdam UMC doctoral fellowship and a Junior Clinical Scientist Fellowship (03-007-2022-0035) from the Dutch Heart Foundation.

