## Supplemental Material for "Polygenic susceptibility to dilated cardiomyopathy underlies peripartum, alcohol-induced, and cancer therapy-related cardiomyopathies"

### Supplementary Materials

#### Supplementary Methods

##### PARTICIPATING STUDIES

###### Mass General Brigham Biobank

The Mass General Brigham (MGB) Biobank is an observational biobank of patients from a multicenter health system in Massachusetts, USA.(1–3) Participants are enrolled with broad-based consent collected by local research coordinators, either as part of a collaborative research study or electronically through a patient portal. Demographic data, blood samples and surveys are collected at baseline and linked to electronic health record data. All adult patients provided informed consent to participate. A small number of children were enrolled with IRB-approved assent forms; upon reaching 18 years of age all enrolled children had to provide consent or were removed from the study. The Human Research Committee of MGB approved the Biobank protocol (2009P002312).

All samples were genotyped using the GlobalScreeningArray version 1. The genotype array data underwent stringent QC. Variant QC consisted of removal of variants with allele count <2, missingness >2%, Hardy Weinberg equilibrium test *P*-value <1e-6 (in each continental super-population), and those with discordant frequencies as compared to gnomAD (X^2 statistic >300; applied in each continental super-population). Sample QC consisted of removal of outliers for heterozygosity or missingness, removal of samples with a mismatch between inferred and self-reported sex, and removal of samples with a mismatch between exome sequencing and array calls. Principal component analysis and relatedness inference were performed using PC-Relate and PC-Air, while ancestry labels (for continental super-populations) were learned from a k-nearest-neighbor model trained on 1000Genomes project data.(4–6) Following these stringent QC procedures, data were subsequently genome-wide imputed to the TOPMed imputation panel (r2) on the Michigan Imputation Server (submitted by batch).(7)

Exome sequencing, as part of NHGRI's *Center for Common Disease Genomics*, has currently been completed for over 53,000 MGB participants. Samples were sequenced on Illumina NovaSeq machines with a custom exome panel (TWIST Human Core Exome), with a target of at least 20X coverage at >85% of target sites.(8) Alignment, processing and joint-calling of variants were performed using the Genome Analysis ToolKit (GATK v4.1) following GATK best practices, after which a stringent QC pipeline was applied to the sequencing data.

Assessments for the presence of hypertension, diabetes mellitus, a family history of DCM, or preeclampsia prior to the development of cardiomyopathies was performed by detailed chart reviews. Chart reviews were also utilized for the identification of the type of cancer and anthracycline agent used for those with CCM. Assessment of antecedent obesity was performed through temporal queries of the MGB biobank to identify individuals with ICD-10 codes E66.01 (morbid obesity due to excess calories) or E66.09 (obesity due to excess calories) prior to or at the time of the addition of the ICD-10 codes for cardiomyopathies to their charts.

**UK Biobank**

The UK Biobank is a large population-based prospective cohort study from the United Kingdom with rich phenotypic and genetic data on 500,000 individuals.(9) Participants were recruited between 2006 and 2010 at ages of 40-69 years. Available genetic data currently include genome-wide imputed data for nearly all participants. Genotyping was performed using Affymetrix UK Biobank Axiom (450,000 samples) and Affymetrix UK BiLEVE axiom (50,000 samples) arrays. The genetic data were subsequently imputed to the Haplotype Reference Consortium panel and UK10K + 1000 Genomes panels (version 3 imputed data). We removed samples that had withdrawn their consent, samples that were outliers for heterozygosity or missingness, individuals with putative sex chromosome aneuploidy, and individuals with a mismatch between self-reported and genetically inferred sex, as determined by central quality-control. The UK Biobank resource was approved by the UK Biobank Research Ethics Committee and all participants provided written informed consent to participate. Use of UK Biobank data was performed under application number 17488 and was approved by the Mass General Brigham Institutional Review Board.

**FinnGen Biobank**

FinnGen, launched in 2017, is a public-private partnership research project that combines genotype data from newly collected and legacy samples administered by Finnish biobanks (https://www.finngen.fi/en) to provide novel insights into human diseases. This study includes genotype data from 453,733 individuals as part of FinnGen Data Freeze 11. The data were linked using unique national personal identification numbers to the national hospital discharge registry (available since 1968), the cause of death registry (since 1969), and the specialist outpatient registry (since 1998).

Newly collected FinnGen samples were genotyped using the FinnGen ThermoFisher Axiom custom array (Thermo Fisher Scientific, San Diego, CA, USA), while legacy cohorts were genotyped using Illumina and Affymetrix arrays (Illumina Inc., San Diego, CA, and Thermo Fisher Scientific, Santa Clara, CA, USA), as previously detailed.(10) Principal component analysis was employed to exclude samples that were not of genotype-inferred Finnish ancestry, following the method described previously for Data Freeze 5 (ref.). Genotype imputation was performed using a population-specific SISu v4 imputation reference panel comprising 8,557 whole genomes, following the protocol available at: https://dx.doi.org/10.17504/protocols.io.xbgfijw.

Data for all FinnGen participants were linked to national registries using unique national personal identification numbers, ensuring comprehensive coverage of health-related outcomes.

**FinnGen Ethics Statement**

FinnGen participants provided informed consent for biobank research, based on the Finnish Biobank Act. Alternatively, separate research cohorts, collected prior to the Finnish Biobank Act came into effect (in September 2013) and start of FinnGen (August 2017), were collected based on study-specific consents and later transferred to the Finnish biobanks after approval by Fimea (Finnish Medicines Agency), the National Supervisory Authority for Welfare and Health. Recruitment protocols followed the biobank protocols approved by Fimea. The Coordinating Ethics Committee of the Hospital District of Helsinki and Uusimaa (HUS) statement number for the FinnGen study is Nr HUS/990/2017.

The FinnGen study is approved by Finnish Institute for Health and Welfare (permit numbers: THL/2031/6.02.00/2017, THL/1101/5.05.00/2017, THL/341/6.02.00/2018, THL/2222/6.02.00/2018, THL/283/6.02.00/2019, THL/1721/5.05.00/2019 and THL/1524/5.05.00/2020), Digital and population data service agency (permit numbers: VRK43431/2017-3, VRK/6909/2018-3, VRK/4415/2019-3), the Social Insurance Institution (permit numbers: KELA 58/522/2017, KELA 131/522/2018, KELA 70/522/2019, KELA 98/522/2019, KELA 134/522/2019, KELA 138/522/2019, KELA 2/522/2020, KELA 16/522/2020), Findata permit numbers THL/2364/14.02/2020, THL/4055/14.06.00/2020, THL/3433/14.06.00/2020, THL/4432/14.06/2020, THL/5189/14.06/2020, THL/5894/14.06.00/2020, THL/6619/14.06.00/2020, THL/209/14.06.00/2021, THL/688/14.06.00/2021, THL/1284/14.06.00/2021, THL/1965/14.06.00/2021, THL/5546/14.02.00/2020, THL/2658/14.06.00/2021, THL/4235/14.06.00/2021, Statistics Finland (permit numbers: TK-53-1041-17 and TK/143/07.03.00/2020 (earlier TK-53-90-20) TK/1735/07.03.00/2021, TK/3112/07.03.00/2021) and Finnish Registry for Kidney Diseases permission/extract from the meeting minutes on 4th July 2019.

The Biobank Access Decisions for FinnGen samples and data utilized in FinnGen Data Freeze 11 include: THL Biobank BB2017_55, BB2017_111, BB2018_19, BB_2018_34, BB_2018_67, BB2018_71, BB2019_7, BB2019_8, BB2019_26, BB2020_1, BB2021_65, Finnish Red Cross Blood Service Biobank 7.12.2017, Helsinki Biobank HUS/359/2017, HUS/248/2020, HUS/430/2021 §28, §29, HUS/150/2022 §12, §13, §14, §15, §16, §17, §18, §23, §58 and §59, Auria Biobank AB17-5154 and amendment #1 (August 17 2020) and amendments BB_2021-0140, BB_2021-0156 (August 26 2021, Feb 2 2022), BB_2021-0169, BB_2021-0179, BB_2021-0161, AB20-5926 and amendment #1 (April 23 2020) and it´s modification (Sep 22 2021), BB_2022-0262, BB_2022-0256, Biobank Borealis of Northern Finland_2017_1013, 2021_5010, 2021_5018, 2021_5015, 2021_5015 Amendment, 2021_5023, 2021_5023 Amendment, 2021_5017, 2022_6001, 2022_6006 Amendment, BB22-0067, 2022_0262, Biobank of Eastern Finland 1186/2018 and amendment 22§/2020, 53§/2021, 13§/2022, 14§/2022, 15§/2022, 27§/2022, 28§/2022, 29§/2022, 33§/2022, 35§/2022, 36§/2022, 37§/2022, 39§/2022, 7§/2023, Finnish Clinical Biobank Tampere MH0004 and amendments (21.02.2020 & 06.10.2020), 8§/2021, 9§/2021, §9/2022, §10/2022, §12/2022, 13§/2022, §20/2022, §21/2022, §22/2022, §23/2022, 28§/2022, 29§/2022, 30§/2022, 31§/2022, 32§/2022, 38§/2022, 40§/2022, 42§/2022, 1§/2023, Central Finland Biobank 1-2017, BB_2021-0161, BB_2021-0169, BB_2021-0179, BB_2021-0170, BB_2022-0256, and Terveystalo Biobank STB 2018001 and amendment 25th Aug

2020, Finnish Hematological Registry and Clinical Biobank decision 18th June 2021, Arctic biobank P0844: ARC_2021_1001.

**FinnGen Acknowledgements**

We acknowledge the participants and investigators of THE FinnGen study. The FinnGen project is funded by two grants from Business Finland (HUS 4685/31/2016 and UH 4386/31/2016) and the following industry partners: AbbVie Inc., AstraZeneca UK Ltd, Biogen MA Inc., Bristol Myers Squibb (and Celgene Corporation & Celgene International II Sàrl), Genentech Inc., Merck Sharp & Dohme LCC, Pfizer Inc., GlaxoSmithKline Intellectual Property Development Ltd., Sanofi US Services Inc., Maze Therapeutics Inc., Janssen Biotech Inc, Novartis Pharma AG, and Boehringer Ingelheim International GmbH. The following biobanks are acknowledged for delivering biobank samples to FinnGen: Auria Biobank (www.auria.fi/biopankki), THL Biobank (www.thl.fi/biobank), Helsinki Biobank (www.helsinginbiopankki.fi), Biobank Borealis of Northern Finland (https://www.ppshp.fi/Tutkimus-ja-opetus/Biopankki/Pages/Biobank-Borealis-briefly-in-English.aspx), Finnish Clinical Biobank Tampere (www.tays.fi/en-US/Research_and_development/Finnish_Clinical_Biobank_Tampere), Biobank of Eastern Finland (www.ita-suomenbiopankki.fi/en), Central Finland Biobank (www.ksshp.fi/fi-FI/Potilaalle/Biopankki), Finnish Red Cross Blood Service Biobank (www.veripalvelu.fi/verenluovutus/biopankkitoiminta), Terveystalo Biobank (www.terveystalo.com/fi/Yritystietoa/Terveystalo-Biopankki/Biopankki/) and Arctic Biobank (https://www.oulu.fi/en/university/faculties-and-units/faculty-medicine/northern-finland-birth-cohorts-and-arctic-biobank). All Finnish Biobanks are members of BBMRI.fi infrastructure (www.bbmri.fi). Finnish Biobank Cooperative -FINBB (https://finbb.fi/) is the coordinator of BBMRI-ERIC operations in Finland. The Finnish biobank data can be accessed through the Fingenious® services (https://site.fingenious.fi/en/) managed by FINBB.

**DCM POLYGENIC SCORE**

A polygenic score for DCM was calculated for each study participant as the weighted sum of the number of effective alleles across the genome, using the Plink2 software. The score was derived from genome-wide association studies (GWAS) of cardiac magnetic resonance imaging-derived measures of left ventricular size and function, specifically focusing on left ventricular end-systolic volume indexed for body surface area (LVESVi), a known quantitative endophenotype for DCM. Polygenic scores derived from LVESVi genetic association data have been shown to effectively predict DCM and have been utilized as such to represent a polygenic liability to DCM.(11,12) Here, a genome-wide polygenic score for DCM was generated from genetic association data on LVESVi using the LDpred2 algorithm, which accounts for linkage disequilibrium across the genome. The final score included 1,137,406 common genetic variants, each with a minor allele frequency greater than 1%. The polygenic score was trained and validated in a subset of the UK Biobank. Importantly, our analysis of secondary cardiomyopathies in the UK Biobank excluded UK Biobank study participants who were part of the polygenic score derivation or training.

**RARE ("MONOGENIC") DCM VARIANTS**

In the MGB Biobank, we leveraged whole exome sequencing data (as described above) to classify carriers of rare pathogenic ("monogenic") variants of relevance to DCM. "Carriers" were defined as study participants harboring a ClinVar pathogenic or likely pathogenic (P/LP) variant from 12 genes with "definitive" or "strong" evidence for DCM pathogenicity, as determined by the international ClinGen consortium: *BAG3*, *DES*, *DSP*, *FLNC*, *LMNA*, *MYH7*, *PLN*, *RBM20*, *SCN5A*, *TNNC1*, *TNNT2*, and *TTN*.(8,13) In addition, for 6 of these genes, we included loss-of-function (LOF) variants based on evidence supporting protein truncation as a mechanism for DCM: *BAG3*, *DSP, FLNC*, *LMNA*, *RBM20*, and *TTN*.(14–19) Relevant LOF variants for *TTN* were restricted to those affecting cardiac isoforms.(13)

Briefly, protein consequences of variants were annotated using dbNSFP (version 4.3a) and the Loss-of-Function Transcript Effect Estimator (LOFTEE) plug-in implemented in the Variant Effect Predictor (VEP; version 105) (https://github.com/konradjk/loftee). VEP was used to determine the most severe consequence of a given variant for each gene transcript. LOFTEE was implemented to identify high-confidence loss-of-function variants (LOF), including frameshift indels, stopgain variants and splice site disrupting variants. LOFs flagged by LOFTEE as dubious were removed, such as LOFs affecting poorly conserved exons and splice variants affecting NAGNAG sites or non-canonical splice regions.

We performed a sensitivity analysis including high-confidence LOF, ClinVar P/LP and missense variants in 91 genes from clinical genetic screening panels for DCM.(19) The following genes were included in this analysis: *ABCC9*, *ACTC1*, *ACTN2*, *AGL*, *ALMS1*, *ALPK3*, *ANKRD1*, *BAG3*, *BRAF*, *CACNA1C*, *CAV3*, *CSRP3*, *DES*, *FLNC*, *FXN*, *GAA*, *HRAS*, *JPH2*, *MYBPC3*, *MYH6*, *MYH7*, *MYL2*, *MYL3*, *MYOZ2*, *PLN*, *PRKAG2*, *PTPN11*, *RAF1*, *RIT1*, *SOS1*, *TCAP*, *TNNC1*, *TNNI3*, *TNNT2*, *TPM1*, *TTR*, *VCL*, *CALR3*, *CASQ2*, *CHRM2*, *CRYAB*, *CTF1*, *CTNNA3*, *DOLK*, *DSC2*, *DSG2*, *DSP*, *DTNA*, *EYA4*, *FHL2*, *FKRP*, *FKTN*, *GATA4*, *GATA6*, *GATAD1*, *HCN4*, *ILK*, *JUP*, *KRAS*, *LAMA4*, *LDB3*, *LMNA*, *LRRC10*, *MAP2K1*, *MAP2K2*, *MIB1*, *MURC*, *MYLK2*, *MYOM1*, *MYPN*, *NEBL*, *NEXN*, *NKX2-5*, *NPPA*, *NRAS*, *PDLIM3*, *PKP2*, *PLEKHM2*, *PRDM16*, *RBM20*, *RYR2*, *SCN5A*, *SGCD*, *SLC22A5*, *TBX20*, *TGFB3*, *TMEM43*, *TMPO*, *TRDN*, *TXNRD2* and *TTN*.

#### Supplementary Results

**Supplementary Table 1: Baseline characteristics in the UK Biobank.** ALCM = alcohol-induced cardiomyopathy; CCM = cancer therapy-induced cardiomyopathy; F = female; PPCM = peripartum cardiomyopathy; SD = standard deviation.

| | PPCM (n=2) | ALCM (n=74) | CCM (n=29) | Control (n=295,055) | P-value |
|---|---|---|---|---|---|
| **Female Gender (%)** | 2 (100.0) | 4 (5.4) | 17 (58.6) | 161,756 (54.8) | <0.001 |
| **Mean age (SD) in years** | 51.5 (0.7) | 58.7 (7.3) | 59.0 (7.3) | 56.5 (8.1) | 0.032 |
| **Race (%)** | | | | | 0.49 |
| **Black** | 0 (0.0) | 3 (4.1) | 2 (6.9) | 5,057 (1.7) | |
| **Chinese** | 0 (0.0) | 0 (0.0) | 0 (0.0) | 998 (0.3) | |
| **Other** | 0 (0.0) | 0 (0.0) | 1 (3.4) | 6,258 (2.1) | |
| **South Asian** | 0 (0.0) | 0 (0.0) | 0 (0.0) | 6,232 (2.1) | |
| **White** | 2 (100.0) | 71 (95.9) | 26 (89.7) | 276,510 (93.7) | |

**Supplementary Table 2: Baseline characteristics in the FinnGen Biobank.** Cigarette smoking data is not available. ALCM = alcohol-induced cardiomyopathy; CCM = cancer therapy-induced cardiomyopathy; F = female; PPCM = peripartum cardiomyopathy; SD = standard deviation.

| | PPCM (n=7) | ALCM (n=125) | CCM (n=65) | Control (n=417,753) | P-value |
|---|---|---|---|---|---|
| **Female Gender (%)** | 7 (100.0) | 12 (9.6) | 43 (66.2) | 241,023 (57.7) | <0.001 |
| **Mean age (SD) in years at onset of disease** | 35.3 (4.3) | 59.2 (11.8) | 56.3 (16.6) | NA | <0.001 |
| **Mean age (SD) in years at death or end of follow-up** | 47.6 (8.7) | 64.6 (10.5) | 64.8 (15.3) | 59.1 (17.7) | <0.001 |
| **Race = White (%)** | 7 (100.0) | 125 (100.0) | 65 (100.0) | 417,753 (100.0) | NA |

**Supplementary Table 3: Presence of clinical risk factors at the time of diagnosis of secondary cardiomyopathy.** ALCM = alcohol-induced cardiomyopathy; CCM = cancer therapy-induced cardiomyopathy; DCM = dilated cardiomyopathy; PPCM = peripartum cardiomyopathy.

| Phenotype | Hypertension | Diabetes mellitus | Family history of DCM | Obesity | Preeclampsia |
|---|---|---|---|---|---|
| **PPCM (%)** | 4 (19.0) | 4 (19.0) | 0 (0) | 2 (9.5) | 5 (23.8) |
| **ALCM (%)** | 47 (61.8) | 20 (26.3) | 5 (6.6) | 9 (11.8) | NA |
| **CCM (%)** | 2 (12.5) | 2 (12.5) | 2 (12.5) | 0 (0) | NA |
| **All secondary cardiomyopathies (%)** | 53 (46.9) | 26 (23.0) | 7 (6.2) | 11 (9.7) | 5 (4.4) |

**Supplementary Table 4: Impact of a DCM polygenic score on chart-validated PPCM, ALCM, and CCM phenotypes in the MGB Biobank.** Odds ratios represent the odds of disease per 1-standard deviation increase in polygenic score. They are reported from logistic regression models including age, sex, race, and the first 10 principal components of genetic ancestry. ALCM = alcohol-induced cardiomyopathy; CCM = cancer therapy-induced cardiomyopathy; CI = confidence interval; OR = odds ratio; PPCM = peripartum cardiomyopathy; SD = standard deviation; SE = standard error.

| | **OR per SD increase in score** | **SE** | **95% CI** | **P-value** |
|---|---|---|---|---|
| **PPCM (n=21)** | 1.78 | 0.23 | 1.13-2.81 | 0.013 |
| **ALCM (n=76)** | 1.99 | 0.12 | 1.57-2.52 | <0.001 |
| **CCM (n=16)** | 1.77 | 0.26 | 1.06-2.98 | 0.03 |

**Supplementary Table 5: Impact of a DCM polygenic score on PPCM, ALCM, and CCM phenotypes in the UK Biobank.** Odds ratios (OR) represent the odds of disease per 1-standard deviation (SD) increase in polygenic score and are reported from logistic regression models including age, sex, race, genotyping array, and the first 10 principal components. ALCM = alcohol-induced cardiomyopathy; CCM = cancer therapy-induced cardiomyopathy; CI = confidence interval; OR = odds ratio; PPCM = peripartum cardiomyopathy; SD = standard deviation; SE = standard error.

| | OR per SD increase in score | SE | 95% CI | P-value |
|---|---|---|---|---|
| **PPCM (n=2)** | 6.71 | 0.82 | 1.35-33.4 | 0.02 |
| **ALCM (n=74)** | 1.16 | 0.12 | 0.92-1.45 | 0.217 |
| **CCM (n=29)** | 1.63 | 0.19 | 1.12-2.37 | 0.01 |

**Supplementary Table 6: Impact of a DCM polygenic score on PPCM, ALCM, and CCM phenotypes in the FinnGen Biobank.** Odds ratios represent the odds of disease per 1-standard deviation increase in polygenic score and are reported from logistic regression models including age, sex, race, genotyping array, and the first 10 principal components. ALCM = alcohol-induced cardiomyopathy; CCM = cancer therapy-induced cardiomyopathy; CI = confidence interval; OR = odds ratio; PPCM = peripartum cardiomyopathy; SD = standard deviation; SE = standard error.

| | **OR per SD increase in score** | **SE** | **95% CI** | **P-value** |
|---|---|---|---|---|
| **PPCM (n=7)** | 1.65 | 0.39 | 0.77-3.54 | 0.2 |
| **ALCM (n=125)** | 1.25 | 0.09 | 1.05-1.49 | 0.014 |
| **CCM (n=65)** | 1.51 | 0.13 | 1.18-1.94 | 0.001 |

**Supplementary Table 7: Risk of PPCM, ALCM, and CCM in individuals with a polygenic score in the top 5th or 20th percentile, in the MGB Biobank.** Odds ratios were calculated by comparing those with a high polygenic score with the remainder of the population in a logistic regression model adjusted for age, sex, race, and the first 10 principal components of ancestry. ALCM = alcohol-induced cardiomyopathy; CCM = cancer therapy-induced cardiomyopathy; CI = confidence interval; OR = odds ratio; PPCM = peripartum cardiomyopathy; SE = standard error.

| Phenotype | Cases | Cases with high polygenic score | OR | SE | CI | P-value |
|---|---|---|---|---|---|---|
| **High polygenic risk cutoff: 95th percentile** | | | | | | |
| PPCM | 21 | 3 | 3.149 | 0.632 | 0.913-10.865 | 0.07 |
| ALCM | 76 | 12 | 4.056 | 0.308 | 2.217-7.421 | <0.001 |
| CCM | 16 | 0 | NA | NA | NA | NA |
| **All secondary cardiomyopathies** | **113** | **113** | **3.339** | **0.272** | **1.958-5.696** | **<0.001** |
| **High polygenic risk cutoff: 80th percentile** | | | | | | |
| PPCM | 21 | 9 | 3.15 | 0.445 | 1.317-7.537 | 0.01 |
| ALCM | 76 | 32 | 2.984 | 0.234 | 1.886-4.72 | <0.001 |
| CCM | 16 | 6 | 2.613 | 0.521 | 0.942-7.25 | 0.065 |
| **All secondary cardiomyopathies** | **113** | **113** | **2.979** | **0.192** | **2.044-4.343** | **<0.001** |

**Supplementary Table 8: Impact of DCM monogenic variant on chart-validated PPCM, ALCM, and CCM phenotypes in the MGB Biobank.** Odds ratios represent the odds of disease among monogenic mutation carriers and are reported from logistic regression models including age, sex, race, and the first 10 principal components. Monogenic variants are composed of ClinVar pathogenic or likely pathogenic (P/LP) variants from 12 genes with "definitive" or "strong" evidence for DCM pathogenicity, as determined by the international ClinGen consortium: *BAG3, DES, DSP, FLNC, LMNA, MYH7, PLN, RBM20, SCN5A, TNNC1, TNNT2,* and *TTN* and curated predicted LOFs for genes where protein truncation is a mechanism for DCM: *BAG3, DSP*, *FLNC*, *LMNA*, *RBM20* and *TTN*. ALCM = alcohol-induced cardiomyopathy; CCM = cancer therapy-induced cardiomyopathy; CI = confidence interval; OR = odds ratio; PPCM = peripartum cardiomyopathy; SD = standard deviation; SE = standard error.

| | OR | SE | 95% CI | P-value |
|---|---|---|---|---|
| **PPCM (n=21)** | 27.21 | 0.79 | 5.80-127.64 | 2.79E-05 |
| **ALCM (n=76)** | 9.51 | 0.53 | 3.39-26.67 | 1.87E-05 |
| **CCM (n=16)** | 13.93 | 1.05 | 1.79-108.60 | 0.012 |

**Supplementary Table 9: Combined polygenic and monogenic assessment of individuals with chart-validated phenotypes in the MGB Biobank.** Results are presented as the number and percentage of individuals divided into 4 classifications of genetic risk: individuals with a high polygenic score only, defined as a score >80th percentile, those with a DCM monogenic variant only, those with both a high polygenic score and a monogenic variant, and individuals with neither a high polygenic score nor a DCM monogenic variant. ALCM = alcohol-induced cardiomyopathy; CCM = cancer therapy-induced cardiomyopathy; PPCM = peripartum cardiomyopathy.

| | Both | High polygenic score only | Monogenic variant only | Neither |
|---|---|---|---|---|
| **PPCM (%)** | 1 (4.8) | 8 (38.1) | 1 (4.8) | 11 (52.4) |
| **ALCM (%)** | 2 (2.6) | 30 (39.5) | 2 (2.6) | 42 (55.3) |
| **CCM (%)** | 1 (6.2) | 5 (31.2) | 0 (0) | 10 (62.5) |
| **Control (%)** | 50 (0.12) | 8,139 (19.82) | 215 (0.52) | 32,652 (79.53) |

**Supplementary Table 10: Brief clinical histories, monogenic variant status, and polygenic score for MGB Biobank participants with peripartum cardiomyopathy.** DCM = dilated cardiomyopathy; Dx = diagnosed; ECMO = extracorporeal membrane oxygenation; F = female; GDMT = guideline-directed medical therapy; HELLP syndrome = hemolysis, elevated liver enzymes, and low platelets syndrome; ICD = implantable cardioverter-defibrillator; IVF = in-vitro fertilization; LGE = late gadolinium enhancement; LV = left ventricle; LVEF = left ventricular ejection fraction; PPCM = peripartum cardiomyopathy; RV = right ventricle; VAD = ventricular assist device; VUS = variant of uncertain significance.

| Case number | Gender | Monogenic variant | Polygenic score | History | Preeclampsia | Hypertension | Diabetes mellitus | Obesity | Family history of DCM |
|---|---|---|---|---|---|---|---|---|---|
| PPCM-1 | F | No | 99 | • Dx in their 50’s after IVF<br>• Complicated by preeclampsia<br>• LVEF 32% and LV dilation, improved to 77% with GDMT, then decreased to 43% after stopping GDMT | Yes | No | No | No | No |
| PPCM-2 | F | No | 98 | • Dx with DCM 6 months of childbirth<br>• Cardiac stress test without evidence of ischemia<br>• LVEF normalized 6 years later, LV dilation persisted | No | Yes | No | Yes | No |
| PPCM-3 | F | No | 96 | • Dx in their 20’s<br>• LVEF 30% postpartum state, improved to 45% | No | Yes | No | Yes | No |
| PPCM-4 | F | No | 93 | • Dx in their 30’s<br>• LVEF 55% pre-pregnancy, dropped to 30% during pregnancy, improved to 55% with GDMT | No | No | No | No | No |

| | | | | | | | | | |
|---|---|---|---|---|---|---|---|---|---|
| PPCM-5 | F | No | 91 | • Dx in their 20's at 6-month postpartum with LVEF 15% and LV dilation<br>• RV biopsy revealed moderate interstitial fibrosis, marked myocyte hypertrophy and no evidence of active myocarditis | No | No | No | No | No |
| PPCM-6 | F | No | 90 | • Dx in their 20's after noting dyspnea 2 weeks postpartum<br>• Prominent LV trabeculation, LVEF 20-25%, improved with GDMT<br>• Clinical genetic testing revealed a *TTN* VUS (heterozygous for a single nucleotide duplication in titin c.30156dupA, p.Tyr10053lefsX3 leading to a frameshift and premature truncation) | No | No | No | No | No |
| PPCM-7 | F | Yes, *TTN* LOF | 87 | • Dx in their 20's postpartum with LVEF 37%, improved to 64% with GDMT<br>• Had a second pregnancy without recurrence of PPCM | No | No | No | No | No |
| PPCM-8 | F | No | 86 | • Dx in their 30's<br>• Pregnancy complicated by preeclampsia/HELLP syndrome<br>• LVEF normalized with GDMT | Yes | No | No | No | No |
| PPCM-9 | F | No | 83 | • Dx in their 40's during a pregnancy<br>• LVEF 40% | No | No | No | No | No |
| PPCM-10 | F | No | 79 | • Dx in their 30's with PPCM after being found to have ventricular arrhythmias during pregnancy | No | No | No | No | No |

| | | | | | | | | | |
|---|---|---|---|---|---|---|---|---|---|
| PPCM-11 | F | No | 74 | • Dx in their late teens after a cardiac post-C-section, LVEF 38%<br>• During second pregnancy, LVEF 65%, decreased to 35% with new global hypokinesis two weeks after delivery, improved to 45% with GDMT | No | No | No | No | No |
| PPCM-12 | F | No | 58 | • Dx in their 20’s in early postpartum period with LVEF 28% from normal during pregnancy (screening echocardiogram due to history of diabetes mellitus)<br>• Cardiac MRI with LVEF 22%, RV ejection fraction 32%, normal perfusion, and no LGE | Yes | Yes | Yes | No | No |
| PPCM-13 | F | Yes, *TTN* LOF | 55 | • Dx in their 20’s<br>• Persistently low LVEF and LV dilation<br>• Required ICD, VAD and heart transplant | No | No | No | No | No |
| PPCM-14 | F | No | 54 | • Dx in their 30’s<br>• LVEF normalized after 3 months | Yes | No | Yes | No | No |
| PPCM-15 | F | No | 50 | • Dx in their 30’s with LVEF 20% during an admission 6 weeks post-partum for heart failure and hypoxic respiratory failure requiring intubation<br>• Etiology felt to be PPCM, although also found to have scleroderma renal crisis<br>• LVEF normalized at subsequent visits | No | No | Yes | No | No |
| PPCM-16 | F | No | 45 | • Referred to Mass General Brigham after PPCM diagnosis<br>• LVEF subsequently normalized with GDMT | No | No | No | No | No |

| | | | | | | | | | |
|---|---|---|---|---|---|---|---|---|---|
| PPCM-17 | F | No | 39 | • Dx in their 30's<br>• LVEF recovered with GDMT | No | No | No | No | No |
| PPCM-18 | F | No | 33 | • Dx in their 30's<br>• LVEF of 20%, partial recovery with GDMT<br>• Had panel testing for DCM which showed 1 variant in GAA for Pompe disease (autosomal recessive), 3 variants of uncertain significance in *MYH7*, *RYR2*, and *TNNI3K* | No | No | No | No | No |
| PPCM-19 | F | No | 29 | • Dx in their 20's at 33 weeks after a cardiac arrest requiring an emergent C-section for twins<br>• LVEF 5-10%<br>• Required ECMO and a heart transplant | No | No | No | No | No |
| PPCM-20 | F | No | 28 | • Dx in their 30's, 1 month post-partum<br>• LVEF of 35-40%<br>• LVEF 55% three years later | No | Yes | Yes | No | No |
| PPCM-21 | F | No | 14 | • Dx with LV dilation but normal LVEF five days after delivery<br>• Also with hepatitis C, polysubstance use, and extensive obstetric history (gravida 10, para 6)<br>• Thought to be PPCM, hypertensive cardiomyopathy or postpartum preeclampsia | Yes | No | No | No | No |

**Supplementary Table 11: Brief clinical histories, monogenic variant status, and polygenic score for MGB Biobank participants with alcohol-induced cardiomyopathy.** ALCM = alcohol-induced cardiomyopathy; ACS = acute coronary syndrome; AF = atrial fibrillation; CAD = coronary artery disease; DCM = dilated cardiomyopathy; F = female; GDMT = guideline-directed medical therapy; HF = heart failure; ICD = implantable cardioverter-defibrillator; LGE = late gadolinium enhancement; LV = left ventricle; LVAD = left ventricular assist device; LVEF = left ventricular ejection fraction; M = male; MRI = magnetic resonance imaging; TICM = tachycardia-induced cardiomyopathy; TTE = transthoracic echocardiogram.

| Case number | Gender | Monogenic variant | Polygenic score | Clinical History | Hypertension | Diabetes mellitus | Obesity | Family history of DCM |
|---|---|---|---|---|---|---|---|---|
| ALCM-1 | M | No | 100 | • LVEF 15%<br>• Heavy alcohol use | Yes | Yes | No | No |
| ALCM-2 | M | No | 100 | • LVEF 19% and dilated LV<br>• Heavy alcohol use | No | No | No | No |
| ALCM-3 | M | No | 100 | • LVEF 15% in their 40's<br>• Heavy alcohol use | Yes | Yes | No | No |
| ALCM-4 | M | No | 100 | • LVEF 15% and dilated LV<br>• Heavy alcohol use<br>• NICM likely secondary to alcohol, could also be secondary to infection or arrhythmia | Yes | Yes | No | No |
| ALCM-5 | M | Yes, *TTN* LOF | 99 | • Biventricular HF, LVEF 26%, dilated LV<br>• Heavy alcohol use<br>• Coronary angiogram only revealed luminal irregularities<br>• Diagnosed with non-ischemic ALCM | Yes | No | No | No |
| ALCM-6 | M | No | 99 | • LVEF 35%<br>• 4 glasses of wine reported daily | No | No | No | No |

| | | | | | | | | |
|---|---|---|---|---|---|---|---|---|
| ALCM-7 | M | No | 99 | • Biventricular HF, LVEF 25%<br>• Heavy alcohol use<br>• ALCM, or less likely ischemic cardiomyopathy | Yes | Yes | No | No |
| ALCM-8 | F | No | 97 | • Admitted with sepsis<br>• TTE with LVEF 16%<br>• Reported >2 drinks/day<br>• Presumed ALCM with possible stress-induced component due to sepsis | Yes | No | No | No |
| ALCM-9 | M | No | 97 | • LVEF 9%, four chamber dilation<br>• AF with heavy alcohol use<br>• ALCM with potential "familial" cardiomyopathy | No | No | No | Yes |
| ALCM-10 | M | No | 97 | • LVEF 26% and dilated LV with diffusely hypokinetic right and left ventricles<br>• Heavy alcohol use | Yes | No | No | No |
| ALCM-11 | M | No | 97 | • LVEF 44% in their 40's<br>• Heavy alcohol use<br>• No other pertinent DCM clinical risk factor | No | No | No | No |
| ALCM-12 | F | No | 96 | • Midrange ejection fraction to 50% with LV dilation<br>• Heavy alcohol use | No | No | Yes | No |
| ALCM-13 | M | No | 95 | • Lowest LVEF noted to be 10% with dilated LV<br>• Heavy alcohol use | Yes | No | No | No |
| ALCM-14 | M | No | 94 | • LVEF 21% and dilated LV<br>• More than 12 drinks daily at the time of diagnosis | Yes | Yes | No | No |
| ALCM-15 | M | No | 93 | • LVEF 10% and dilated LV<br>• Heavy alcohol use | No | No | No | No |

| | | | | | | | | |
|---|---|---|---|---|---|---|---|---|
| ALCM-16 | M | No | 93 | • LVEF 22% and dilated LV<br>• Heavy alcohol use | No | No | No | No |
| ALCM-17 | M | No | 92 | • LVEF 26% and dilated LV<br>• Heavy alcohol use | Yes | No | No | No |
| ALCM-18 | M | No | 91 | • LVEF 40%<br>• Heavy alcohol use<br>• Thought to be ALCM | Yes | No | Yes | No |
| ALCM-19 | M | No | 90 | • LVEF 33% and dilated LV<br>• 5 drinks of hard liquor daily at the time of diagnosis<br>• Father had DCM | No | No | No | Yes |
| ALCM-20 | M | No | 90 | • LVEF 45% and dilated LV<br>• Heavy alcohol use | Yes | No | No | No |
| ALCM-21 | M | No | 89 | • LVEF 15% and dilated LV<br>• Long-standing heavy alcohol use | Yes | No | No | No |
| ALCM-22 | M | Yes, *TTN* LOF | 88 | • HF in their 20's<br>• Heavy alcohol use<br>• Required a heart transplant 10 years later | No | No | No | No |
| ALCM-23 | M | No | 88 | • LVEF 20%<br>• Heavy alcohol use<br>• Some reports of cocaine use | No | Yes | No | No |
| ALCM-24 | M | No | 87 | • HF in their 40's<br>• Heavy alcohol use<br>• Exercise myocardial perfusion imaging showed scar in left anterior descending artery territory without evidence of ischemia | Yes | No | No | No |

| | | | | | | | | |
|---|---|---|---|---|---|---|---|---|
| ALCM-25 | M | No | 87 | • HF with a history of CAD but no known ACS preceding the diagnosis<br>• Chronic heavy alcohol use | Yes | No | No | No |
| ALCM-26 | F | No | 87 | • HF with midrange ejection fraction at 48%<br>• Heavy alcohol use<br>• Cardiac MRI with minimal mid-myocardial LGE<br>• Amount of enhancement was minimal; ALCM most likely diagnosis | Yes | No | No | No |
| ALCM-27 | M | No | 86 | • LVEF 23% and dilated LV<br>• HF specialist thought HF secondary to heavy alcohol use and possible genetic predisposition due to family history of DCM<br>• GDMT initiated, LVEF normalized | Yes | No | No | Yes |
| ALCM-28 | M | No | 85 | • LVEF 16% and dilated LV with diffuse hypokinesis<br>• Heavy alcohol use | Yes | Yes | No | No |
| ALCM-29 | M | No | 85 | • LVEF 34% and dilated LV with global hypokinesis<br>• Heavy alcohol use<br>• GDMT initiated, alcohol abstinence, LVEF normalized | No | No | No | No |
| ALCM-30 | M | No | 84 | • LVEF 15%<br>• Heavy alcohol use<br>• Reports of cocaine use | Yes | Yes | No | No |
| ALCM-31 | M | No | 82 | • LVEF 30%<br>• Heavy alcohol use<br>• Ischemic evaluation unrevealing<br>• Thought to be ALCM | Yes | No | No | No |
| ALCM-32 | F | No | 82 | • Lowest LVEF 30% and dilated LV | No | No | No | No |

| | | | | | | | | |
|---|---|---|---|---|---|---|---|---|
| | | | | • Heavy alcohol use and cirrhosis | | | | |
| ALCM-33 | M | No | 77 | • LVEF 45%<br>• Heavy alcohol use | No | No | No | No |
| ALCM-34 | F | No | 76 | • LVEF 26%<br>• Heavy alcohol use | Yes | No | Yes | No |
| ALCM-35 | M | No | 75 | • LVEF 39%<br>• Heavy alcohol use and disulfiram use for alcohol use disorder<br>• Coronary angiogram with only a minor plaque found in the left anterior descending artery<br>• Cardiac MRI without evidence of myocardial inflammation or scarring | Yes | No | No | No |
| ALCM-36 | F | No | 74 | • Long-standing DCM, lowest LVEF 14% and dilated LV cavity<br>• Heavy alcohol use<br>• Thought to be ALCM or to a viral cardiomyopathy | Yes | Yes | No | No |
| ALCM-37 | F | No | 74 | • LVEF 40% and dilated LV cavity with global hypokinesis in their 30's<br>• Heavy alcohol use and cirrhosis | No | No | No | No |
| ALCM-38 | M | No | 74 | • LVEF 20%<br>• Heavy alcohol use | No | No | No | No |
| ALCM-39 | F | No | 70 | • LVEF 15% in their 40's<br>• 6 beers daily | Yes | No | No | No |
| ALCM-40 | M | No | 70 | • LVEF 45% and dilated LV<br>• Heavy alcohol use | Yes | No | Yes | No |
| ALCM-41 | M | No | 69 | • Chronic DCM, lowest LVEF 16% and dilated LV | No | Yes | No | Yes |

| | | | | | | | | |
|---|---|---|---|---|---|---|---|---|
| | | | | cavity<br>• Heavy alcohol use<br>• Recurrent congestive HF exacerbations thought to be secondary to alcohol use (relapse) and GDMT non-adherence<br>• Nephew had DCM | | | | |
| ALCM-42 | M | No | 69 | • LVEF 30% and normal LV cavity size with increased LV wall thickness<br>• Heavy alcohol use<br>• ALCM or secondary to iron infiltration | No | No | No | No |
| ALCM-43 | M | No | 69 | • LVEF 15% and dilated LV<br>• Heavy alcohol use<br>• ALCM most likely diagnosis by providers | Yes | No | No | No |
| ALCM-44 | F | No | 68 | • LVEF 10% and dilated LV cavity<br>• Heavy alcohol use<br>• ALCM most likely diagnosis by providers | Yes | Yes | Yes | No |
| ALCM-45 | M | No | 68 | • LVEF 10%, required temporary LVAD with heavy alcohol use<br>• Coronary angiogram without CAD<br>• GDMT initiated, alcohol abstinence, LVEF improved to 52% | No | Yes | No | No |
| ALCM-46 | M | No | 68 | • LVEF 15% and dilated LV cavity<br>• Heavy alcohol use | No | No | No | No |
| ALCM-47 | M | No | 65 | • LVEF 40%, normal LV cavity size<br>• Heavy alcohol use, cirrhosis, pancreatitis and peripheral neuropathy | No | No | No | No |
| ALCM-48 | M | No | 64 | • LVEF 45% | No | Yes | Yes | No |

| | | | | | | | | |
|---|---|---|---|---|---|---|---|---|
| | | | | • Heavy alcohol use<br>• ALCM most likely diagnosis by providers | | | | |
| ALCM-49 | M | No | 63 | • LVEF 30% and LV cavity dilation<br>• Heavy alcohol use<br>• ALCM most likely diagnosis by providers | Yes | Yes | No | No |
| ALCM-50 | M | No | 62 | • LVEF 23%, dilated LV cavity; also with tetralogy of Fallot, underwent repair<br>• Heavy alcohol use<br>• Cardiac stress test unrevealing | No | No | No | No |
| ALCM-51 | M | Yes, *TTN* LOF | 61 | • LVEF 21% with dilated LV<br>• Heavy alcohol use<br>• Required a heart transplant<br>• Brother with DCM of unknown etiology, and father with presumed ischemic cardiomyopathy | Yes | No | No | Yes |
| ALCM-52 | M | No | 61 | • LVEF 15%<br>• Heavy alcohol use | Yes | No | Yes | No |
| ALCM-53 | F | No | 60 | • LVEF 10% with dilated LV<br>• Coronary angiogram unrevealing<br>• ALCM most likely diagnosis by providers | Yes | No | No | No |
| ALCM-54 | M | No | 60 | • LVEF 19% with dilated LV<br>• Heavy alcohol use | Yes | Yes | No | No |

| | | | | | | | | |
|---|---|---|---|---|---|---|---|---|
| ALCM-55 | M | No | 59 | • LVEF 46% and normal LV cavity size<br>• Heavy alcohol use<br>• Besides ALCM, other potential etiologies were thought to be ischemic cardiomyopathy (due to coronary calcifications on chest imaging) or secondary to hemochromatosis<br>• Stress cardiac MRI without evidence of obstructive plaque or hemochromatosis, ALCM felt to be the most likely etiology | No | No | No | No |
| ALCM-56 | M | No | 58 | • LVEF 23% with dilated LV; advanced age on diagnosis<br>• Heavy alcohol use<br>• Thought to be ALCM or ischemic cardiomyopathy | Yes | Yes | No | No |
| ALCM-57 | M | No | 58 | • LVEF 15% with dilated LV<br>• Heavy alcohol use | No | No | No | No |
| ALCM-58 | M | No | 57 | • LVEF 14% with dilated LV<br>• Heavy alcohol use | Yes | No | No | No |
| ALCM-59 | M | No | 57 | • LVEF 35%, normal LV<br>• Heavy alcohol use | Yes | No | Yes | No |
| ALCM-60 | M | No | 54 | • LVEF 15% with dilated LV cavity<br>• 5 beers daily | Yes | Yes | No | No |
| ALCM-61 | F | No | 51 | • LVEF 10% with dilated LV<br>• Heavy alcohol use<br>• Coronary angiogram unrevealing, presumed ALCM | Yes | No | No | No |
| ALCM-62 | F | No | 51 | • ALCM per chart review, no further information available in the chart as the patient was transferred to the Mayo Clinic for a cardiac | No | No | No | No |

| | | | | | | | | |
|---|---|---|---|---|---|---|---|---|
| | | | | transplant. | | | | |
| ALCM-63 | M | No | 39 | • Presented to the hospital with alcohol withdrawal syndrome, had symptoms of HF, TTE with LVEF 50% with normal LV size and diffuse hypokinesis | No | No | No | No |
| ALCM-64 | M | No | 34 | • LVEF 14% with normal LV cavity<br>• Heavy alcohol use | Yes | No | No | No |
| ALCM-65 | M | No | 33 | • LVEF 20% with dilated LV<br>• 400 milliliters of whisky daily | Yes | No | No | No |
| ALCM-66 | M | No | 32 | • LVEF 13% with normal LV cavity<br>• One pint of vodka daily | Yes | Yes | No | No |
| ALCM-67 | M | No | 31 | • LVEF 44% with normal LV cavity size<br>• 6 beers daily; also with cirrhosis | Yes | No | No | No |
| ALCM-68 | F | No | 26 | • LVEF 10% with dilated LV<br>• Heavy alcohol use; also with a remote history of doxorubicin therapy<br>• Coronary angiogram unrevealing<br>• Most likely ALCM as supported by improvement with alcohol cessation | No | No | No | No |
| ALCM-69 | M | No | 25 | • LVEF 38% with dilated LV with increased wall thickness<br>• Chronic daily alcohol use but no heavy use (2 alcoholic drinks daily) | Yes | Yes | No | No |

| | | | | | | | | |
|---|---|---|---|---|---|---|---|---|
| ALCM-70 | M | No | 24 | • LVEF 20% with dilated LV<br>• Heavy alcohol use; also with tobacco and cocaine use<br>• Stress test unrevealing, therefore assumed to have non-ischemic DCM secondary to alcohol and/or cocaine use | Yes | Yes | Yes | No |
| ALCM-71 | M | Yes, *TTN* LOF | 17 | • LVEF 17% with dilated LV<br>• 1.5 liters of liquor daily on weekends<br>• Thought to be ALCM | No | No | No | No |
| ALCM-72 | M | No | 16 | • Long-standing HF<br>• Heavy alcohol use | Yes | No | No | No |
| ALCM-73 | M | No | 14 | • LVEF 40% with dilated LV<br>• Heavy alcohol use | Yes | No | No | No |
| ALCM-74 | M | No | 13 | • LVEF 20%<br>• 6 to 10 beers daily | Yes | No | No | No |
| ALCM-75 | M | No | 7 | • LVEF 16% with normal LV cavity size<br>• Heavy alcohol use<br>• Required ICD placement<br>• GDMT initiated, alcohol abstinence, LVEF improved to 50% | Yes | No | No | No |
| ALCM-76 | M | No | 5 | • Diagnosed with ALCM remotely, without much information available in the chart<br>• ICD placed after an episode of ventricular tachycardia storm | No | No | No | No |

**Supplementary Table 12: Brief clinical histories, monogenic variant status, and polygenic score for MGB Biobank participants with cancer therapy-induced cardiomyopathy.** CCM = cancer therapy-induced cardiomyopathy; DCM = dilated cardiomyopathy; F = female; HF = heart failure; LVEF = left ventricular ejection fraction; M = male; MRI = magnetic resonance imaging; TTE = transthoracic echocardiogram.

| Case number | Gender | Monogenic variant | Polygenic score | Clinical History | Cardiotoxic chemo agent(s) | Malignancy | Hypertension | Diabetes mellitus | Obesity | Family history of DCM |
|---|---|---|---|---|---|---|---|---|---|---|
| CCM-1 | F | No | 94 | • LVEF 66% to 50% on surveillance TTE after the fourth cycle of anthracyclines | Anthracycline and trastuzumab | Invasive ductal carcinoma of the breast | No | No | No | No |
| CCM-2 | F | Yes, *TTN* LOF | 94 | • HF 15 years after completion of 4 cycles of anthracyclines and radiation to the left breast<br>• Ischemic and non-ischemic evaluations unrevealing | Doxorubicin | Ductal carcinoma of the breast | No | No | No | No |
| CCM-3 | F | No | 88 | • LVEF 45-50% after completion of chemotherapy with anthracyclines and trastuzumab | Doxorubicin and trastuzumab | Breast cancer | No | No | No | No |
| CCM-4 | F | No | 87 | • LVEF 32% 10 years after treatment with anthracyclines | Doxorubicin | Breast cancer | No | No | No | No |
| CCM-5 | M | No | 83 | • Moderately reduced LVEF to 49% 10 years after completing a treatment with anthracyclines | Doxorubicin | Osteosarcoma | No | No | No | No |

| | | | | | | | | | | |
|---|---|---|---|---|---|---|---|---|---|---|
| | | | | • Total doxorubicin dose totaled 375 milligrams per square meter<br>• GDMT initiated, LVEF improved to 60% | | | | | | |
| CCM-6 | F | No | 83 | • HF with midrange ejection fraction on surveillance TTE after treatment with anthracyclines<br>• Lowest LVEF 46%, improved with GDMT | Doxorubicin | Osteosarcoma | No | No | No | No |
| CCM-7 | M | No | 79 | • LVEF from 51% prior to chemotherapy with anthracyclines to 23% six years later<br>• Father had DCM | Doxorubicin | Diffuse large cell B-cell lymphoma | No | Yes | No | Yes |
| CCM-8 | M | No | 73 | • LVEF from 63% prior to chemotherapy to 12% seven years after anthracycline therapy<br>• Received a total of 90 mg per square meter of doxorubicin<br>• Required a cardiac transplant | Doxorubicin | Osteosarcoma | No | No | No | No |
| CCM-9 | F | No | 73 | • LVEF from 57% prior to chemotherapy to 30% three years after treatment with anthracyclines<br>• Lifetime doxorubicin dose of 500 milligrams per square | Doxorubicin | Non-Hodgkin's lymphoma | No | No | No | No |
| CCM-10 | M | No | 65 | • LVEF 20% months after completing anthracycline therapy | Doxorubicin | Diffuse large cell B-cell lymphoma | No | No | No | No |

| | | | | | | | | | | |
|---|---|---|---|---|---|---|---|---|---|---|
| | | | | • No prior baseline TTE<br>• GDMT initiated, LVEF improved to 50% | | | | | | |
| CCM-11 | M | No | 52 | • LVEF from 52% prior to chemotherapy to 29% four years after doxorubicin therapy<br>• Ischemic workup unrevealing, suggesting CCM | Doxorubicin | Diffuse large cell B-cell lymphoma | No | No | No | No |
| CCM-12 | M | No | 49 | • TTE with LVEF 20-25%<br>• LVEF improved to 45-50% after initiation of chemotherapy<br>• A few months after receiving anthracyclines, LVEF 35% | Doxorubicin | Anaplastic large cell lymphoma | No | No | No | No |
| CCM-13 | F | No | 44 | • LVEF 17% and biventricular dysfunction 6 years after receiving doxorubicin<br>• Serologies for autoimmune or infiltrative pathologies, coronary angiogram and cardiac MRI were unrevealing | Doxorubicin | Invasive ductal carcinoma of the breast | Yes | No | No | No |
| CCM-14 | M | No | 33 | • LVEF from 55% to 48% while receiving chemotherapy | Doxorubicin | Mediastinal large B-cell lymphoma | No | No | No | No |
| CCM-15 | F | No | 32 | • LVEF from 60% prior to chemotherapy to 35% two years after treatment with anthracyclines | Doxorubicin | Diffuse large B-cell lymphoma | Yes | Yes | No | Yes |
| CCM-16 | M | No | 28 | • LVEF from 60% prior to chemotherapy to 40% five | Daunorubicin | Acute myelocytic | No | No | No | No |

| | | | | | | |
|---|---|---|---|---|---|---|
| | | | | months after anthracycline therapy<br>• Lowest LVEF 25%<br>• Ischemic workup unrevealing | | leukemia |
